# Adults’ self-reported barriers and enablers to riding a bike for transport: a systematic review

**DOI:** 10.1101/2022.04.22.22274159

**Authors:** Lauren Pearson, Danielle Berkovic, Sandy Reeder, Belinda Gabbe, Ben Beck

## Abstract

Riding a bike for transport purposes is an effective way to improve population and environmental health. Despite this, participation levels in many countries are low. Identifying the barriers and enablers to riding a bike for transport is essential to developing interventions that encourage bike riding. In this mixed-methods systematic review, we aimed to identify the perceived barriers and enablers to adults riding a bike for transport in Organisation for Economic Development (OECD) countries. A systematic database search was conducted to identify relevant peer-reviewed and grey literature. Fifty-five papers/reports met eligibility criteria. There were 34 barriers and 21 enablers identified. The leading barriers related to riding on the road alongside motor vehicles. Other factors identified included the provision and quality of cycling infrastructure, personal factors such as physical fitness, attitudinal factors such as community perceptions of cyclists, and environmental factors. While this review highlights the complexity of factors that influence the uptake of riding a bike for transport, many of the leading factors could be overcome through the provision of high-quality protected infrastructure for bike riders. Other interventions to address other known barriers and enablers are needed to increase the uptake of bike riding.

## Introduction

The health benefits of riding a bike are vast and well established, including improved cardiorespiratory fitness, cognitive function and mental wellbeing (Celis-Morales et al., 2017; Leyland et al., 2019; Nordengen et al., 2019). Beyond this, an increase in trips made by bike has the potential to reduce traffic congestion, environmental pollutants, and offer economic benefits to both individuals and governments (Litman, 2015; Pérez et al., 2017; Rabl & De Nazelle, 2012). A large proportion of trips made by car are a distance that could potentially be made by bike (Beck, 2021; Department of Transport, 2021; Harper, 2021; Vagane, 2007), highlighting the potential for increased participation in bike riding for transport, in addition to for recreational purposes.

Despite the benefits of bike riding, and increased accessibility from e-bikes (Fishman & Cherry, 2016; Fyhri et al., 2017), moving away from car-based transport and towards bike riding has become a considerable cultural, behavioural and political challenge (Agarwal & Gupta, 2021; Kent, 2015; Richards et al., 2011; Wilson & Mitra, 2020). Recognising what facilitates, and what prevents people from choosing to cycle for transport permits informed decisions for cycling infrastructure, promotion campaigns and policy reform for government and advocacy organisations.

There are several studies that report the barriers and enablers of riding a bike for transport in local populations, cited regularly throughout active transport-related literature. However, these study findings are often extrapolated to populations with potentially different circumstances. Further, there are a variety of measures and populations reported, limiting the potential for comparison between studies.

There is, at present, no peer-reviewed systematic review that synthesises the findings of, and appraises the quality of studies, from multiple contexts of the barriers and enablers to riding a bike for transport. This review is important because it allows for identification of potential issues in measurement of barriers and enablers, areas of needed research, and consistencies and differences between countries.

The aim of this systematic review was to identify the perceived barriers and enablers to adults’ participation in riding a bike for transport. The inclusion of qualitative research in this systematic review attempted to represent some of these lived experiences while still maintaining large-scale quantitative studies to facilitate generalisability and reproducibility.

## Methods

Systematic searches of databased of peer-reviewed scientific and grey literature were undertaken to identify the barriers and enablers of riding a bike for transport in Organisation for Economic Co-operation and Development (OECD) countries (OECD, 2020). Both quantitative and qualitative studies were included, and no publication year limits were applied due to the limited published synthesis of data prior to this review. Trips made by bike exclusively for transport purposes were chosen as they have likely differing barriers and enablers than when trips are made for recreation. Transport included any journey made to travel to a destination, such as commuting and to travel to shops. Most OECD member countries are high-income and developed economies (Canser et al., 2016). Because of this, they share similar cycling infrastructure, laws and volumes of traffic (Buehler & Pucher, 2017). Due to these similarities, only OECD countries were included in this review to allow for meaningful comparisons. The review included papers published up to the 5^th^ February, 2021, when the search was conducted.

### Eligibility criteria

Primary studies published in a peer-reviewed journal or grey literature were eligible for inclusion if they:

- reported on either the perceived barriers or enablers of riding a bike for transport purposes (including initiating participation, or riding more than they currently do),
- recruited an adult population,
- were published in English and;
- were conducted in an OECD country.

Bikes were defined as manual pushbikes and electric bikes. Where trip purpose was not defined in full-text, the paper was excluded. Perceived barriers and enablers included factors that participants reported as either preventing them or encouraging them to ride a bike hypothetically.

To enable findings between homogenous study types to be synthesised, we focussed on studies that stated self-reported barriers and/or enablers to cycling. Studies that aimed to elucidate potential behaviours based on a selection of factors (such as stated preference surveys) or studies that modelled associations between self-reported barriers and frequency of cycling, without reporting the aggregate self-reported barriers and enablers data for the sample, were excluded.

### Peer-reviewed Literature

Peer-reviewed published articles were systematically searched in Ovid Medline, Ovid PsycINFO, Embase, Scopus, Transport Research Information Database (TRID) and EBSCO CINAHL using terms generated through thorough consultation with a librarian and text mining. Text mining involved identifying sentinel papers on the research question and using keywords and title words to develop a list of terms to include in the search. These combined various components including active transport or bicycling and barriers, enablers, attitudes and behaviours. The full list of search terms are shown in Supplementary Materials D. The online platform ‘Covidence’ (Covidence, 2020) was used to store articles imported from databases and facilitate organised and blinded screening. Two independent reviewers screened results first by title and abstract, followed by full-text for adherence to inclusion and exclusion criteria. Conflicts between reviewers about which studies to include were discussed by reviewer one and two until consensus was reached, or if this was not possible a third reviewer was consulted.

Following article screening, a hand-search of the reference list of included studies was conducted by a study author to identify further relevant literature.

### Grey Literature

Research regarding bike riding is often conducted by government and bicycling advocacy organisations. For this reason, a grey literature search was conducted as this type of research is often not peer-reviewed. The term “barriers and enablers of cycling” was systematically searched in Google, Analysis and Policy Observatory (APO), TRID and the OECD Library. An Advanced Search was performed using Google and limiting the search criteria to each OECD country. Through consultation with a librarian specialist in grey literature searching, it was decided that the first 50 results produced by Google would be used. Reviewer one conducted these searches and screen-captured the results, before compiling them into a document. Each result in the document was then screened by title and description (where available) by reviewer one and two. Reviewers were blinded to each other’s results until completion. The method used in resolving conflicts in the peer-reviewed literature was repeated.

### Quality Assessment

The Joanna Briggs Critical Appraisal Tools were used to assess the methodological rigour of peer-reviewed literature (Munn et al., 2014). Tools were specific to study design and included closed questions regarding the methodology, recruitment, analysis and conclusions of the study. Possible answers included ‘yes’, ‘no’, ‘unclear’ or ‘not applicable’.

Reviewers one and two conducted quality assessment independently for each peer-reviewed article after full-text screening. When complete, results of quality assessments were compared. Where there were differences of more than 20%, reviewers discussed until consensus or consulted with reviewer three. A mean appraisal score was then calculated for each article based on the proportion of quality appraisal checklist items met by the study. Articles scoring 50% or below on the mean appraisal score were not included in the review to ensure only high-quality literature was synthesised.

Grey literature was appraised using the Authority, Accuracy, Coverage, Objectivity, Date & Significance (AACODS) Checklist (Tyndall, 2010), a critical appraisal tool specific to grey literature. The AACODS Checklist comprises appraisal of literature based on the accuracy, authority, coverage, objectivity, date and significance. The appraisal process used in the peer-reviewed literature assessment was repeated for the grey literature.

### Data Extraction

Data were extracted on study and participant characteristics and study findings (barriers and enablers). An iterative process was employed to categorise barriers and enablers into broader themes. Outcome measures were extracted for quantitative studies, and participation characteristic and quotes extracted from qualitative studies.

Where data were reported as the proportion of the sample that agreed that a particular factor was a barrier or enabler (binary outcome), the population sample size and outcome were extracted for inclusion in a meta-analysis. Due to the variability in outcome measures, meta-analyses of other data types beyond binary outcomes were not possible. To be included in the meta-analysis, study questions, study populations and outcome measures needed to be comparable. Meta-analyses were conducted in RStudio (RStudio Team, 2020) with the ‘meta’ package (Schwarzer & Schwarzer, 2012). Due to the diversity of populations included in the meta-analyses, random effects models were most suited. Where heterogeneity was high, a restricted maximum likelihood indicator was used.

### Synthesis

A segregated approach was used to synthesise quantitative and qualitative data (Lizarondo L, 2020). Quantitative data were extracted into a table and meta-analysis performed where possible. If meta-analyses were not possible, data were extracted into a separate table for inclusion in supplementary material. Qualitative data were extracted into a table, analysed through coding and quotes presented to illustrate higher level categories of codes. Findings of qualitative and quantitative analyses were reported alongside each other where findings overlapped.

#### Quantitative data

All quantitative data regarding study characteristics were extracted into a table. Outcome data were extracted into two separate tables exclusively for either barriers or enablers. If data were presented stratified into groups, overall cohort data were extracted.

#### Qualitative data

Quotes extracted from qualitative studies were analysed with an inductive approach of thematic coding as described by Gibbs (Gibbs, 2007). All quotes were retrieved from included articles that met selection criteria. Quotes were coded with the same label where they were an example of the same phenomenon, idea or explanation. Codes were organised to provide meaning and structure to the data, leading to higher level categories based on their relationships with each other. To describe each of these categories, quotes were used in results. Analysis was conducted by two reviewers for credibility and rigour.

## Results

There were 4,602 records screened, of which 60 studies met inclusion criteria (Figure 1). To be included in the meta-analysis, study questions walking and cycling data being aggregated, the study not reporting on barriers and/or enablers, and the study reporting only on actual barriers and enablers that were calculated from self-reports and infrastructure than surrounded the participant, or from self-reports and their frequency of cycling.

**Figure 1.**
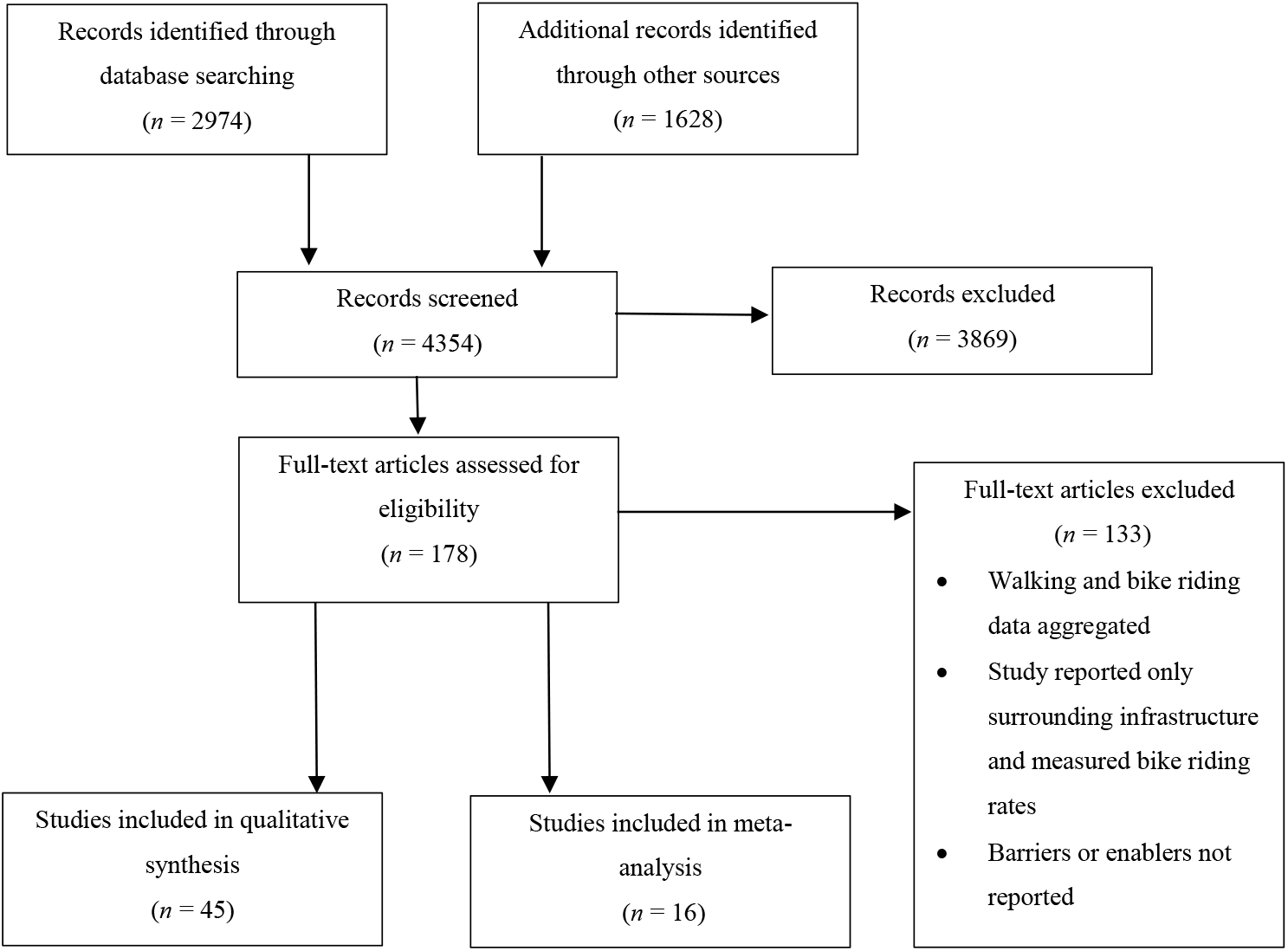
Summary of number of studied included in each screening process

Through quality assessment, 7 publications and 2 grey literature reports did not meet the requirements. Reasons included lack of information about participants and recruitment, and lack of clarity in the measurement and reporting of outcomes (see Supplementary Materials A).

The final 45 final studies were mostly quantitative and cross-sectional in design (*n* = 29), followed by qualitative (*n* = 8), mixed methods (*n* = 7) and one longitudinal survey (see Table 1). Thirty-seven studies were identified from a bibliographic database search, seven from grey literature sources and one from an integrated database (TRID). Online surveys were the most common data collection method (*n* = 23). The majority of studies reported on manual bikes (*n* = 43), and two on electric pedal-assisted bikes (e-bikes). A range of methods were used to assess barriers and enablers and quantify their relative importance, including binary outcomes, Likert scales and other types of proportional data.

**Table 1.** Included study characteristics and cohort description

| **First Author** | **Country**<br>**Study**<br>**Conducted In** | **Sample Size (*n*)** | **Population** | **Study Design** |
| --- | --- | --- | --- | --- |
| Abasahl et al. (2018) | USA | 698 | University staff and students | Cross-sectional survey |
| Agarwal & North (2012) | USA | 103 | University students | Cross-sectional survey |
| Akar et al. (2013) | USA | 2012 | University staff and students | Cross-sectional survey |
| Chowdhury & Costello (2016) | New Zealand | 1178 | Representative sample of Auckland's population - cyclists and non-cyclists after the installation of new cycleways | Cross-sectional survey |
| Community Vibe on behalf of Central Goldfields Shire Council (2017) | Australia | 100 | Community members of a regional area in Victoria, Australia | Mixed-methods |
| Crawford et al. (2001) | Scotland | 690 | Two workplaces within Glasgow - one university and one hospital | Cross-sectional survey |
| Cred Consulting on behalf of City of Sydney and C40 | Australia | 89 | Women who ride a bike for a substantial part of their transport and live in Sydney, Australia | Mixed-methods* |
| Curto et al. (2016) | Spain | 814 | Commuters living and working in Barcelona who are able to ride a bike for 20 minutes | Cross-sectional survey |
| Daley et al. (2007) | Australia | 70 | Men and women recruited through phoning a number on a flyer | Qualitative |
| de Cunha & da Silva (2018) | Portugal | 505 | Staff and PhD students at the Faculty of Engineering, University of Porto (Portugal) | Cross-sectional survey |
| de Geus et al. (2008) | Belgium | 343 | Working Flemish adults who lived within 10km of their workplace | Cross-sectional survey |
| Delso et al. (2018) | Spain | 3786 | Sample of 3786 adults who completed the Vitoria-Gasteiz household travel survey | Quantitative |
| Dill & McNeil (2016) | USA | 3116 | Sample of 3000 adults who lived in the 50 largest U.S. metropolitan areas | Quantitative |
| Fernández-Heredia et al. (2014) | Spain | 3048 | University staff and students | Cross-sectional survey |
| Galway et al. (2021) | Canada | 30 | Thirty adult cyclists living in North America | Qualitative |
| Heart Foundation on behalf of the Cycling Promotion Fund (2011) | Australia | 515 | Random sample of 1000 Australian adults | Cross-sectional survey |
| Heart Foundation on behalf of the Cycling Promotion Fund (2013) | Australia | 1007 | Adult women mostly residing in urban areas in Australia | Cross-sectional survey |
| Heesch et al. (2012) | Australia | 1862 | Members of a community bicycle organisation in Queensland | Cross-sectional survey |
| Heinen et al. (2011) | Netherlands | 4299 | Employees from several large companies in cities in the Netherlands | Cross-sectional survey |
| Ingunn Stangeby on behalf of Transportøkonomisk institutt (1997) | Norway | 392 | Mostly middle-aged people with a high level of education and high household income | Cross-sectional survey |
| Iwinska et al. (2018) | Poland | 561 | Random sample for quantitative survey was people at home in Warsaw during a door to door survey. The cyclist survey was of people who indicated they were cyclists in a previous travel survey, living in Warsaw. Qualitative methods were used for cyclists, transportation activists and a scientific expert on transportation systems in Warsaw. | Mixed-methods |
| Jones et al. (2016) | Netherlands & England | 22 | E-bike owners living in Randstaf or Groningen in the Netherlands and in Oxford in the UK | Qualitative |
| Lechaud (2016) | Portugal | 305 | Convenience sample of 701 people living in Portugal | Cross-sectional survey |
| Leger et al. (2019) | Canada | 37 | Older people living in retirement homes, in the community or in active lifestyle communities | Qualitative |
| Lois et al. (2016) | Spain | 21 | People who commute for work or study in two cities in Spain (Madrid or Vitoria-Gasteiz) | Qualitative |
| Manaugh et al. (2016) | Canada | 4944** | Staff and students at the McGill University in Montreal | Cross-sectional survey |
| McManus et al. (2005) | Australia | 1657 | People who attended a bike to work breakfast event in the CBD of Perth, Western Australia | Cross-sectional survey |
| Mullan (2013) | Ireland | 16 | Leisure and sport cyclists living in Ireland who participated in a non-competitive cycling event (Sean Kelly tour) | Qualitative |
| Munoz et al. (2013) | Spain | 224 | People walking in the centre of Madrid who were local to the area | Cross-sectional survey |
| Nikitas (2018) | Greece | 640 | Sample of adults living in Drama, Greece | Cross-sectional survey |
| Rérat (2019) | Switzerland | 13700 | People who participated in the Bike to Work campaign in Switzerland | Mixed-methods |
| Sahlqvist & Heesch (2012) | Australia | 183 | Members of a community bicycle organisation in Queensland | Cross-sectional survey |
| Salvo et al. (2018) | Canada | 14 | People who have recently relocated to a different neighbourhood | Mixed-methods |
| Schneider & Hu (2015) | USA | 3566 | University of Wisconsin - Milwaukee staff and students who lived away from campus | Cross-sectional survey |
| Scott (2009) | Australia | 300 | Infrequent/non-cyclists living in New South Wales, Australia | Mixed-methods |
| Sears et al. (2012) | USA | 185 | Group of working adults in Vermont who commute by bicycle two or more miles each way on 28 specified days in a 10-month period | Longitudinal survey |
| Štastná et al. (2018) | Czech Republic | 383 | Students and staff of the Faculty of AgriSciences, Mendel University in Brno | Cross-sectional survey |
| Swiers et al. (2017) | England | 194 | University students living in England | Cross-sectional survey |
| Ton et al. (2019) | Netherlands | 2425 | Census data from the Netherlands Mobility Panel. Mostly representative of the Dutch population except for teenagers and low-income individuals | Cross-sectional survey |
| van Bekkum et al. (2011) | Scotland | 831 | Staff and PhD students in a cycle-friendly building in a university in Edinburgh | Cross-sectional survey |
| van Bekkum et al. (2011) | Scotland | 15 | Cycle commuters and potential cycle commuters from a "cycle-friendly" workplace in a Scottish city (Edinburgh) | Qualitative |
| Whannell et al. (2011) | Australia | 270 | First year undergraduate students studying a science, technology and society course focussed on environmental and sustainability issues | Cross-sectional survey |
| Winters et al. (2011) | Canada | 1402 | Current and potential cyclists in Vancouver from a random sample of home addresses | Cross-sectional survey |
| Winters et al. (2015) | Canada | Survey = 191<br>Interviews = 27 | Older adults living in downtown Vancouver, Canada | Mixed-methods |
| Zander et al. (2013) | Australia | 17 | Adults living in inner west Sydney between 50 and 75 who were willing to cycle two hours per week or more for a 12-week period | Qualitative |

\*quantitative outcome data extracted from cross-sectional survey only
\*\*barriers recorded for $n = 295$

### Quantitative measures

There were 16 different outcome measures used across 36 quantitative studies of barriers and enablers. Of these, most studies (*n* = 17) reported the proportion of people who noted that the factor was a barrier or enabler to them as a binary outcome of “yes” or “no”. Remaining quantitative studies used Likert scales (*n* = 9) or other types of proportional measures. Likert scales had further variety, ranging from -1 to 1, to 1 to 7 scales, measuring the importance of a barrier or enabler, level of agreement that it was a barrier or enabler, or how much the factor affected their propensity to ride a bike. Studies that used Likert scale measures reported the mean for the cohort. Other types of measures used included the proportion of people who “somewhat agreed” or “strongly agreed” that a particular factor was a barrier or enabler, the proportion of days not commuted by bike due to a particular barrier, the proportion of people who selected a particular factor as being the main barrier to them riding a bike, and the proportion that reported an enabling factor as the most important improvement for them to start using a bike.

### Participants

Included studies were conducted in Europe (*n* = 21, 48%), North America (*n* = 12, 27%), Australia (*n* = 11, 23%) and New Zealand (*n* = 1, <1%). Quantitative study sample sizes ranged from *n* = 89 to *n* = 13,700, with a median (quartile 1, quartile 3) of 665 (285, 1937) participants. Studies had varying proportions of women (range: 20-100%, median (quartile 1, quartile 3) = 50% (42%, 58%). Eleven of the 45 included studies focussed on university student and/or staff populations exclusively.

### Barriers

There were 34 barriers to cycling for transport identified through quantitative and qualitative methods (Table 2). The most commonly reported and measured barriers in both quantitative and qualitative studies were bad weather, the time taken to a destination being too great, a perceived lack of safety and high density of motor vehicles. Barriers reported in exclusively in qualitative data that were not measured in quantitative studies included a lack of connectivity of bike paths, negative non-rider attitudes toward cyclists, and negative perceptions of e-bikes from people riding pedal bikes. Barriers were categorised into eight themes; safety, trip factors, personal factors, infrastructure, access, environmental factors, end of trip facilities and perceptions. All barriers identified through data that could not be meta-analysed are shown in Supplementary materials B.

**Table 2.** Summary of barriers reported in quantitative and qualitative studies

| Barrier | Qualitative studies (n) | Quantitative studies (n) | Total (n) |
| --- | --- | --- | --- |
| *Safety* |  |  |  |
| Fear of motorist aggression | 4 | 6 | 10 |
| Perceived risk of injury | 2 | 10 | 12 |
| High traffic density | 7 | 10 | 17 |
| Feelings of safety | 3 | 14 | 17 |
| Fear of theft or crime | 4 | 6 | 10 |
| *Infrastructure* |  |  |  |
| Poor quality and condition of dedicated bike lanes | 5 | 6 | 11 |
| Poor condition of roads | 3 | 6 | 9 |
| Limited dedicated bike lanes | 5 | 8 | 13 |
| Darkness | 2 | 8 | 10 |
| Lack of connectivity of bike paths | 3 | 0 | 3 |
| *Trip factors* |  |  |  |
| Lack of storage on bike | 3 | 8 | 11 |
| Long distance | 1 | 11 | 12 |
| Time to destination too great | 4 | 14 | 18 |
| Children to transport | 4 | 11 | 15 |
| *Personal factors* |  |  |  |
| Having to change or shower at destination | 6 | 6 | 12 |
| Too much effort | 2 | 7 | 9 |
| Lack of fitness | 2 | 6 | 8 |
| Not interested | 0 | 6 | 6 |
| Unable to ride a bike due to injury or disability | 1 | 7 | 8 |
| Lack of knowledge of bike riding road rules and bike maintenance | 4 | 6 | 10 |
| Do not own suitable clothing | 0 | 4 | 4 |
| Lack of comfort | 1 | 3 | 4 |
| *Access* |  |  |  |
| No bike | 0 | 8 | 8 |
| Cost of bike too high | 2 | 7 | 9 |
| Mandatory helmet laws | 1 | 1 | 2 |
| *Environmental factors* |  |  |  |
| Bad weather | 3 | 22 | 25 |
| Air-pollution exposure | 2 | 8 | 10 |
| Too many hills | 1 | 8 | 9 |
| *End of trip facilities* |  |  |  |
| Lack of showers or lockers | 0 | 13 | 13 |
| Lack of bike parking | 4 | 12 | 16 |
| *Perceptions* |  |  |  |
| Negative community perception of cyclists | 5 | 0 | 5 |
| Negative perception of e-bikes from pedal cyclists | 2 | 0 | 2 |

Twenty-six barriers met the inclusion criteria for meta-analyses. The summary estimate and corresponding confidence intervals for each outcome are displayed in Figure 2.

**Figure 2.**
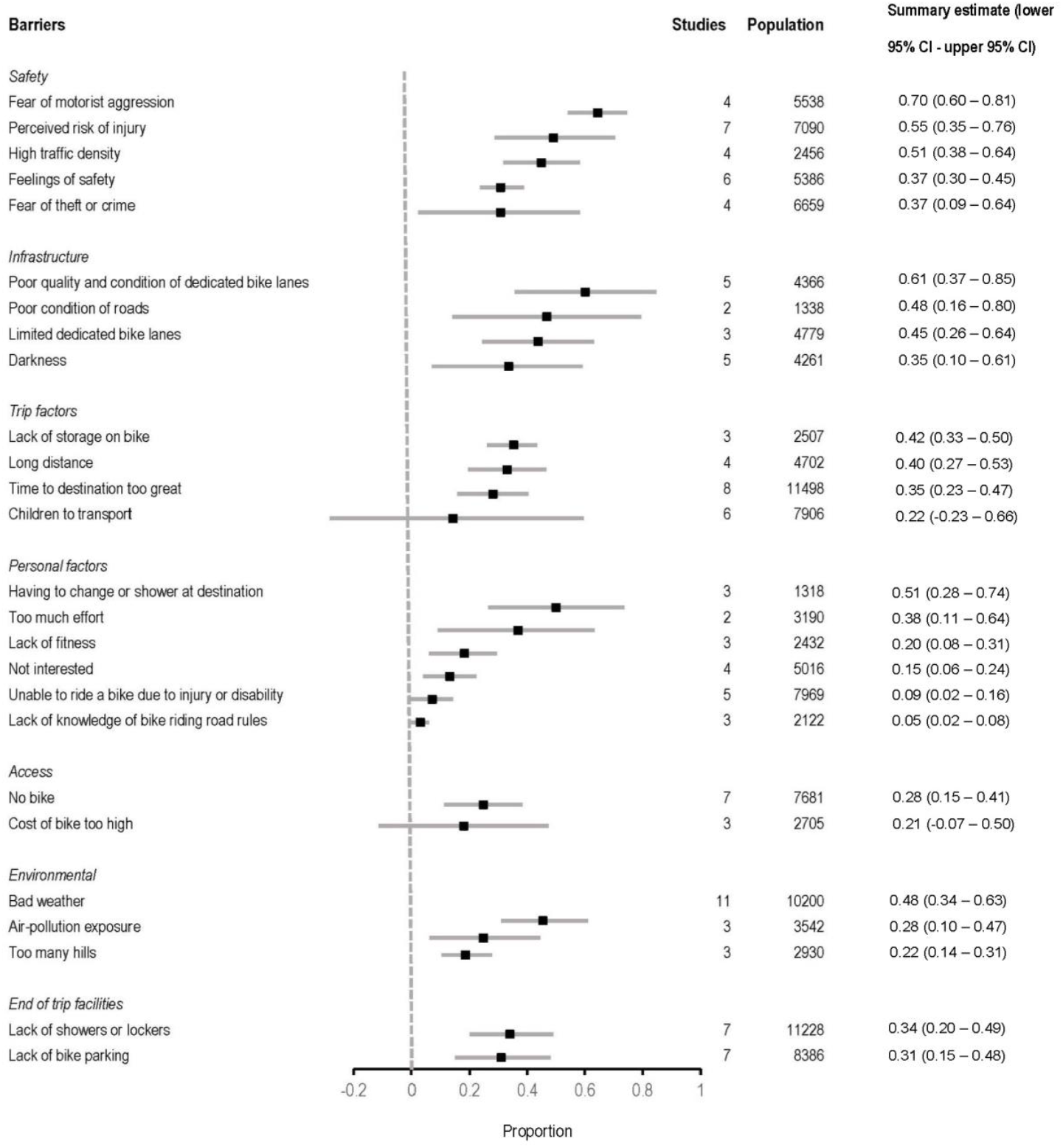
Forest plot of meta-analysis summary estimates (square) and confidence intervals (grey error bars) of the proportion of people who reported each barrier to cycling for transport

#### Safety

Five barriers were reported across 31 studies that related to safety (see Table 3). These included perceived risk of injury, traffic density, feeling of safety, fear of motorist aggression and fear of crime. A fear of motorist aggression was reported by the largest summary estimate relative to other barriers that were meta-analysed (summary estimate = 70%; 95% CI: 60%; 80%). This result was supported by Likert scale data, where participants rated barriers relating to safety as important and qualitative data where participants commented on not feeling safe on a bike (see Supplementary Materials B).

**Table 3.** Outcome table of safety, infrastructure and trip factor-related barriers

| Author, Year | Safety |  |  |  |  | Infrastructure |  |  |  |  | Trip factors |  |  |  |  |  |
| --- | --- | --- | --- | --- | --- | --- | --- | --- | --- | --- | --- | --- | --- | --- | --- | --- |
|  | Fear of motorist aggression | Perceived risk of injury | High traffic density | Feelings of safety | Fear of theft or crime | Poor quality and condition of dedicated bike lanes | Poor condition of roads | Limited dedicated bike lanes | Darkness | Lack of connectivity of bike paths | Lack of storage on bike | Long distance | Time to destination too great | Children to transport | Other activities before/after work | Need vehicle for work |
| Abasahl et al. (2018) | X | X | X |  | X | X | X |  | X |  | X |  | X |  |  |  |
| Agarwal & North (2012) |  |  | X |  |  |  |  |  | X |  |  |  |  |  |  |  |
| Akar et al. (2013) |  |  | X |  |  |  |  |  | X |  |  |  |  |  |  |  |
| Chowdhury (2016) | X |  |  |  |  |  |  | X | X |  | X | X | X | X |  |  |
| Community Vibe (2017) |  |  | X | X |  |  |  | X |  |  |  |  | X |  |  |  |
| Crawford et al. (2001) |  |  | X |  |  |  | X |  |  |  |  | X | X | X |  |  |
| Curto et al. (2016) |  |  |  | X |  |  |  |  |  |  | X |  |  | X |  |  |
| Daley et al. (2007) | X |  |  |  |  | X | X | X |  | X |  |  |  |  |  |  |
| de Geus et al. (2008) |  |  |  |  |  |  |  |  |  |  |  |  | X |  |  |  |
| Delso et al. (2018) |  |  | X | X |  |  |  | X |  |  |  | X |  |  |  |  |
| Dill & McNeil (2016) |  |  | X | X |  |  |  | X |  |  |  | X |  |  |  | X |
| Fernández-Heredia (2014) |  |  |  | X | X |  |  |  |  |  |  | X |  |  |  |  |
| Heart Foundation (2011) |  | X | X |  |  |  | X | X |  |  |  | X | X | X |  |  |
| Heart Foundation (2013) | X | X | X |  |  |  |  |  |  |  |  |  | X | X |  |  |
| Heesch et al. (2012) | X |  | X |  |  |  |  |  |  |  |  | X | X |  |  |  |
| Iwinska et al. (2018) |  | X |  |  |  | X |  | X |  |  |  |  |  |  |  |  |
| Jones et al. (2016) |  |  |  | X |  | X |  |  |  |  |  |  |  | X |  |  |
| Lechaud (2016) |  |  |  | X |  | X |  | X |  |  |  |  |  |  |  |  |
| Leger et al. (2019) |  |  | X |  |  |  |  |  |  |  |  |  |  |  |  |  |
| Lois et al. (2016) | X | X |  | X | X |  |  |  | X |  | X |  | X | X |  |  |
| Manaugh et al. (2016) |  |  |  | X |  |  |  |  |  |  |  | X |  |  |  |  |
| Mullan (2013) |  |  |  |  |  | X |  | X |  | X |  | X |  |  |  |  |
| Nikitas (2018) |  |  |  | X |  | X |  |  |  |  |  |  |  |  |  |  |
| Rérat (2019) |  | X | X |  | X | X |  | X | X |  | X |  | X | X | X |  |
| Sahlqvist & Heesch (2012) | X |  | X |  |  |  |  |  |  |  |  | X |  |  |  |  |
| Salvo et al. (2018) |  |  |  |  |  | X |  |  |  |  |  |  |  |  |  |  |
| Schneider & Hu (2015) |  |  |  | X | X |  | X |  |  |  |  | X | X |  |  |  |
| Scott (2009) | X | X | X | X | X |  | X | X |  | X |  |  | X | X |  | X |
| Sears et al. (2012) |  |  |  |  |  |  |  |  | X |  |  |  |  |  | X |  |
| Štastná et al. (2018) |  | X |  |  | X |  |  |  |  |  |  |  |  |  |  |  |
| Swiers et al. (2017) |  |  |  | X |  |  |  |  |  |  |  |  |  |  |  |  |
| van Bekkum et al. (2011) |  |  |  | X |  |  | X |  | X |  |  | X | X |  |  |  |
| van Bekkum et al. (2011) | X |  | X |  |  | X |  |  |  |  | X |  | X |  |  |  |
| Whannell et al. (2011) |  |  |  | X |  |  |  |  | X |  |  | X |  |  |  |  |
| Winters et al. (2011) | X | X | X |  |  |  | X |  | X |  |  |  |  |  |  |  |
| Winters et al. (2015) |  |  | X |  | X |  |  | X |  |  |  |  |  |  |  |  |
| Zander et al. (2013) |  |  | X |  |  |  |  |  |  |  |  |  |  |  |  |  |

> “I mean the big resistance to cycling is…a lot of people just feel…don’t feel safe cycling” (Jones et al., 2016)

Feeling unsafe was another barrier relating to safety measured in 16 studies (Table 3). When asked about feelings of safety, some participants believed this was regarding their safety on the road in relation to other road-users, or safety on the bike. Many reported feeling unsafe riding on the road due to the presence of motor vehicle traffic and the potential for injury.

> “I don’t feel safe at the moment, I know a few people, parents or whatever who have been hit by or killed by cars in bike accidents… you are not protected like you are in a car” (Jones et al., 2016)

Other participants reported feeling unsafe on a bike due to potential crime.

> “It is dangerous to cycle in the city, except for certain areas” Male, non-cyclist (Scott, 2009)
>
> “I work late and I don’t feel safe. In a car you can lock the doors, but on a bike you just have to ride fast.” (Scott, 2009)

#### Infrastructure

A lack of dedicated bike infrastructure had a higher summary estimate relative to other barriers (summary estimate = 45%; 95% CI: 26%; 64%). This was supported by participant reports across five qualitative studies (M. Daley et al., 2007; Jones et al., 2016; Mullan, 2013; Rérat, 2019; Salvo et al., 2018), and quantitative studies reporting proportional data not included in the meta-analyses (Jennifer Dill & McNeil, 2016; Léchaud, 2016).

Ten studies reported specifically on the poor condition of existing bike infrastructure as a barrier (Table 3), with a high summary estimate relative to all meta-analyses conducted (summary estimate = 61%; 95% CI: 37%; 85%). This was further expanded in qualitative studies, where participants had varied concerns regarding the condition of infrastructure, including; paths not being wide enough to accommodate bikes (M. Daley et al., 2007), bike lanes being adjacent to parked cars where they were at risk of being hit by a door (Winters et al., 2015), lanes being blocked by parked cars (M. Daley et al., 2007), and poor physical condition of bike infrastructure, including from potholes, gravel and litter (Galway et al., 2021; Jones et al., 2016; Rérat, 2019). While not measured in quantitative studies, a lack of connectivity of existing bike infrastructure was reported across four qualitative studies (M. Daley et al., 2007; Linden et al., 2020; Mullan, 2013; Scott, 2009).

> “You are going along the road and all of a sudden there’s a pedestrian crossing or something like that, and the cycle lanes just ends, and then you have to work your way back out into traffic.” (Mullan, 2013)

#### Trip factors

A lack of storage on a bike had the highest summary estimate compared to other trip factors reported (summary estimate = 42%; 95% CI: 33%; 50%). Findings varied greatly across meta-analysis results, Likert scale and qualitative data for other barriers relating to trip factors. Of the five studies that used Likert scales to measure agreement or importance of distance as a barrier, participants in three studies (Crawford et al., 2001; Delso et al., 2018; Fernández-Heredia et al., 2014) reported that this was important or stopped them from cycling, while two studies (Manaugh et al., 2017; Jennifer E. van Bekkum et al., 2011) reported that it was not important or disagreed.

Results varied between studies reporting having children or other people to transport as a barrier (Table 3). Of the two studies that used Likert scales, one reported having people to transport as an important barrier (Crawford et al., 2001), while another found it not discouraging (Jennifer E. van Bekkum et al., 2011). Participants in some qualitative studies felt infrastructure was not available for them to safely transport their children by bike.

> “I would not ride a bike with a small kid in a trailer or child seat on the main roads…It is too dangerous in my mind, even though I always ride on main roads when I am alone” (Rérat, 2019)

#### Personal Factors

Ten personal barriers were reported across 25 studies (Table 4). Having to change or shower at a destination had the largest summary estimate relative to other personal factors meta-analyses (summary estimate = 51%; 95% CI: 28%; 74%). This was supported by findings from four qualitative studies (David Lois et al., 2016; Mullan, 2013; Scott, 2009; J. E. van Bekkum et al., 2011), however accounted for only 0.7% of days not biked for this reason in a longitudinal study of barriers to cycle-commuting (Sears et al., 2012).

**Table 4.**
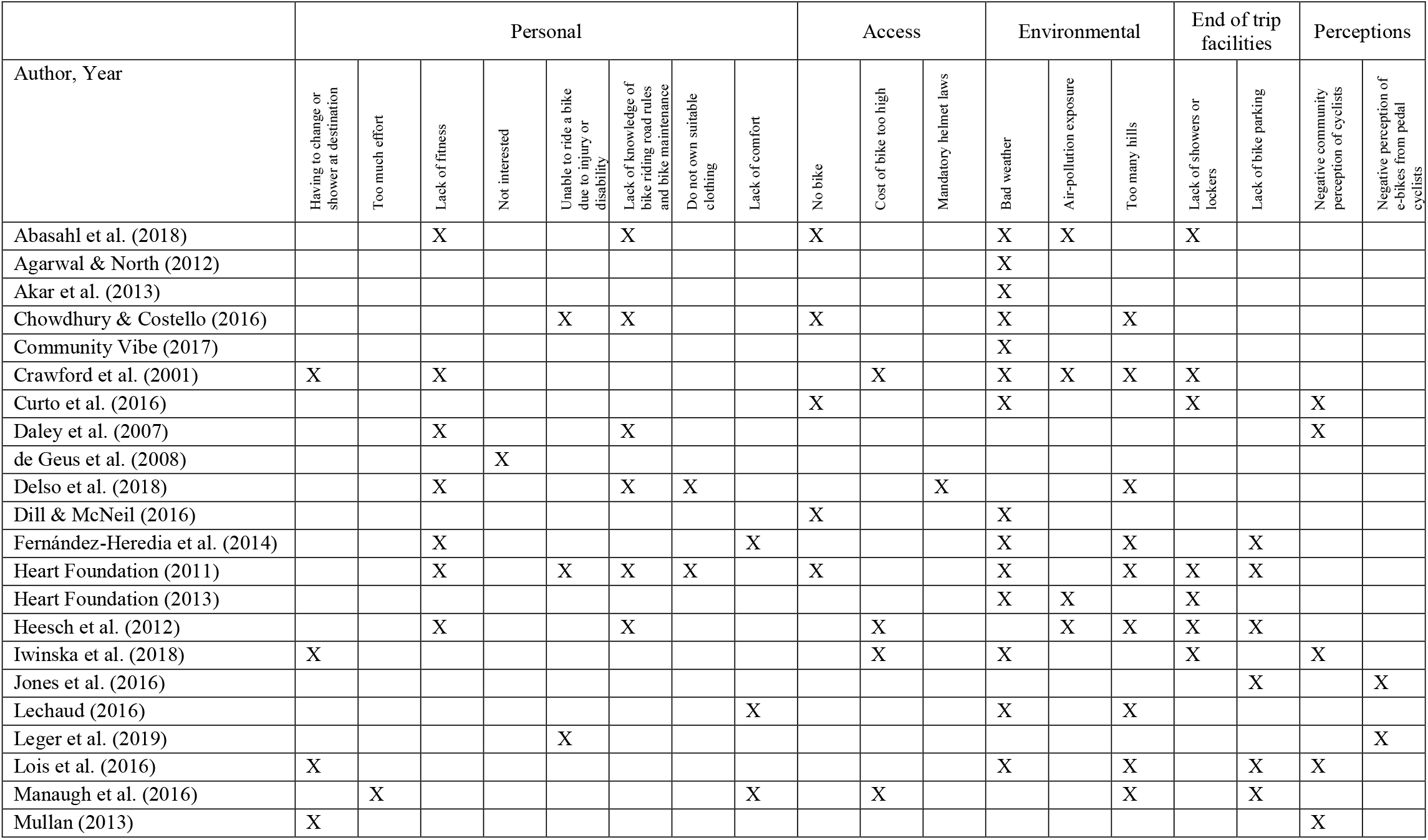
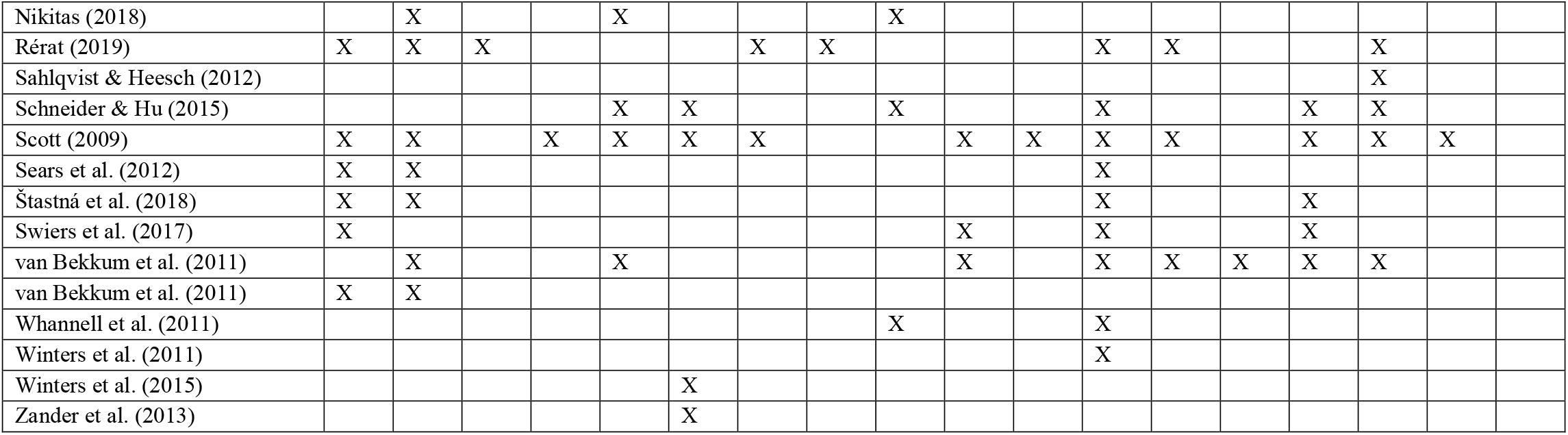
Outcome table of personal, access, environmental, end of trip facility and perception-related barriers

|  | Personal |  |  |  |  |  |  |  | Access |  |  | Environmental |  |  | End of trip facilities |  | Perceptions |  |
| --- | --- | --- | --- | --- | --- | --- | --- | --- | --- | --- | --- | --- | --- | --- | --- | --- | --- | --- |
| Author, Year | Having to change or shower at destination | Too much effort | Lack of fitness | Not interested | Unable to ride a bike due to injury or disability | Lack of knowledge of bike riding road rules and bike maintenance | Do not own suitable clothing | Lack of comfort | No bike | Cost of bike too high | Mandatory helmet laws | Bad weather | Air-pollution exposure | Too many hills | Lack of showers or lockers | Lack of bike parking | Negative community perception of cyclists | Negative perception of e-bikes from pedal cyclists |
| Abasahl et al. (2018) |  |  | X |  |  | X |  |  | X |  |  | X | X |  | X |  |  |  |
| Agarwal & North (2012) |  |  |  |  |  |  |  |  |  |  |  | X |  |  |  |  |  |  |
| Akar et al. (2013) |  |  |  |  |  |  |  |  |  |  |  | X |  |  |  |  |  |  |
| Chowdhury & Costello (2016) |  |  |  |  | X | X |  |  | X |  |  | X |  | X |  |  |  |  |
| Community Vibe (2017) |  |  |  |  |  |  |  |  |  |  |  | X |  |  |  |  |  |  |
| Crawford et al. (2001) | X |  | X |  |  |  |  |  |  | X |  | X | X | X | X |  |  |  |
| Curto et al. (2016) |  |  |  |  |  |  |  |  | X |  |  | X |  |  | X |  | X |  |
| Daley et al. (2007) |  |  | X |  |  | X |  |  |  |  |  |  |  |  |  |  | X |  |
| de Geus et al. (2008) |  |  |  | X |  |  |  |  |  |  |  |  |  |  |  |  |  |  |
| Delso et al. (2018) |  |  | X |  |  | X | X |  |  |  | X |  |  | X |  |  |  |  |
| Dill & McNeil (2016) |  |  |  |  |  |  |  |  | X |  |  | X |  |  |  |  |  |  |
| Fernández-Heredia et al. (2014) |  |  | X |  |  |  |  | X |  |  |  | X |  | X |  | X |  |  |
| Heart Foundation (2011) |  |  | X |  | X | X | X |  | X |  |  | X |  | X | X | X |  |  |
| Heart Foundation (2013) |  |  |  |  |  |  |  |  |  |  |  | X | X |  | X |  |  |  |
| Heesch et al. (2012) |  |  | X |  |  | X |  |  |  | X |  |  | X | X | X | X |  |  |
| Iwinska et al. (2018) | X |  |  |  |  |  |  |  |  | X |  | X |  |  | X |  | X |  |
| Jones et al. (2016) |  |  |  |  |  |  |  |  |  |  |  |  |  |  |  | X |  | X |
| Lechaud (2016) |  |  |  |  |  |  |  | X |  |  |  | X |  | X |  |  |  |  |
| Leger et al. (2019) |  |  |  |  | X |  |  |  |  |  |  |  |  |  |  |  |  | X |
| Lois et al. (2016) | X |  |  |  |  |  |  |  |  |  |  | X |  | X |  | X | X |  |
| Manaugh et al. (2016) |  | X |  |  |  |  |  | X |  | X |  |  |  | X |  | X |  |  |
| Mullan (2013) | X |  |  |  |  |  |  |  |  |  |  |  |  |  |  |  | X |  |
| Nikitas (2018) |  | X |  |  | X |  |  |  | X |  |  |  |  |  |  |  |  |  |
| Rérat (2019) | X | X | X |  |  |  | X | X |  |  |  | X | X |  |  | X |  |  |
| Sahlqvist & Heesch (2012) |  |  |  |  |  |  |  |  |  |  |  |  |  |  |  | X |  |  |
| Schneider & Hu (2015) |  |  |  |  | X | X |  |  | X |  |  | X |  |  | X | X |  |  |
| Scott (2009) | X | X |  | X | X | X | X |  |  | X | X | X | X |  | X | X | X |  |
| Sears et al. (2012) | X | X |  |  |  |  |  |  |  |  |  | X |  |  |  |  |  |  |
| Štastná et al. (2018) | X | X |  |  |  |  |  |  |  |  |  | X |  |  | X |  |  |  |
| Swiers et al. (2017) | X |  |  |  |  |  |  |  |  | X |  | X |  |  | X |  |  |  |
| van Bekkum et al. (2011) |  | X |  |  | X |  |  |  |  | X |  | X | X | X | X | X |  |  |
| van Bekkum et al. (2011) | X | X |  |  |  |  |  |  |  |  |  |  |  |  |  |  |  |  |
| Whannell et al. (2011) |  |  |  |  |  |  |  |  | X |  |  | X |  |  |  |  |  |  |
| Winters et al. (2011) |  |  |  |  |  |  |  |  |  |  |  | X |  |  |  |  |  |  |
| Winters et al. (2015) |  |  |  |  |  | X |  |  |  |  |  |  |  |  |  |  |  |  |
| Zander et al. (2013) |  |  |  |  |  | X |  |  |  |  |  |  |  |  |  |  |  |  |

> “I would have to be in my gear - my cycle pants - and you know, I would have to have [a shower], my hair would be stuck to my head. In my line of work, it’s all about being professional…how you appear, like. If I got caught in the rain it would take from my confidence and take from my work” (Mullan, 2013)

Owning clothes that are suitable to ride a bike in was reported by five cross-sectional studies, with differing findings (Table 4). Proportions of participants who either slightly or strongly agreed that this was a barrier ranged from 19-24% (Léchaud, 2016; Rérat, 2019). Between 5 and 7.6% of participants in two quantitative cross-sectional studies reported not owning suitable clothes as their main barrier to cycling (Heart Foundation, 2013; Scott, 2009). However, on a Likert scale, a survey of women at a university in Edinburgh reported this as not a discouraging factor (Jennifer E. van Bekkum et al., 2011).

#### Access

Access-related barriers were reported in 15 studies. The summary estimate for not owning a bike being a barrier was lower relative to other meta-analyses (Figure 2). The cost of a bike was reported in eight studies (Table 4), where of three using Likert scales, two reported cost as not being a discouraging factor (Manaugh et al., 2017; Jennifer E. van Bekkum et al., 2011) while one reported this as an important barrier (Crawford et al., 2001).

> “My biggest barrier is the cost of a bike” (Swiers et al., 2017)

#### Environmental factors

Environmental barriers were reported in 30 studies. Bad weather was identified as a barrier in in 22 papers (Table 4), with the largest summary estimate for environmental barriers (summary estimate = 48%; 95% CI: 34%; 63%). Consistent with these findings, participants in studies using Likert scales commonly reported bad weather as an important barrier (Crawford et al., 2001; Fernández-Heredia et al., 2014; Jennifer E. van Bekkum et al., 2011; Winters et al., 2011). Some participants in qualitative studies reported that bad weather could put them at risk of injury.

> “It’s not getting wet, but the fact that [in the rain] I can’t see well, and especially at night, I think car drivers will have trouble seeing me and braking and reacting” (David Lois et al., 2016)

#### End of Trip Facilities

A lack good quality bike parking, had a lower summary estimate relative to other meta-analyses (summary estimate = 31%; 95% CI: 15%; 48%). Across four qualitative studies, and two Likert scale studies (Fernández-Heredia et al., 2014; Manaugh et al., 2017) participants reported this as a barrier (Jones et al., 2016; D. Lois et al., 2016; Rérat, 2019; Scott, 2009). One study that reported Likert scales and sampled only women reported this as not a discouraging factor for riding a bike (Jennifer E. van Bekkum et al., 2011).

#### Attitudes

All attitudinal barriers were identified through qualitative data only. Participants reported the negative community perceptions of cyclists in five studies (Table 4). Reasons behind these perceptions differed, including the perception of status when riding a bike, and the perception of cyclists on roads. Some participants reported that when they cycled on the road, they felt that they were treated as a “second-class citizen” (M. Daley et al., 2007). Others reported that if someone were riding a bike, this may lead them to thinking the person does not have a car. They reported having a car as being a mark of socioeconomic status.

> “There is that attitude that you only cycle because you can’t afford a car” (Male, cyclist) (Mullan, 2013)

Negative perceptions of e-bikes from pedal cyclists were reported in two qualitative studies (Jones et al., 2016; S. J. Leger et al., 2019). Participants reported riding an e-bike as being perceived as “cheating” by people who ride unpowered pedal bikes.

> “I guess the initial reaction is that it’s cheating, partly because I’m part of the cycling group/culture and [they] think it’s cheating” (Jones et al., 2016)

### Enablers

There were 22 enablers to cycling for transport identified (Table 5). Enablers were categorised as either relating to infrastructure, resources and end of trip facilities, personal improvement and enjoyment, motivations and practicality. Riding a bike being a fun enjoyable activity was reported by the highest number of studies (*n* = 17), followed by riding a bike being an environmental choice (*n* = 15) and an efficient mode of transport (*n* = 17). All recorded qualitative and quantitative data that were not able to be included in meta-analyses are shown in Supplementary Materials or available on request.

**Table 5.**
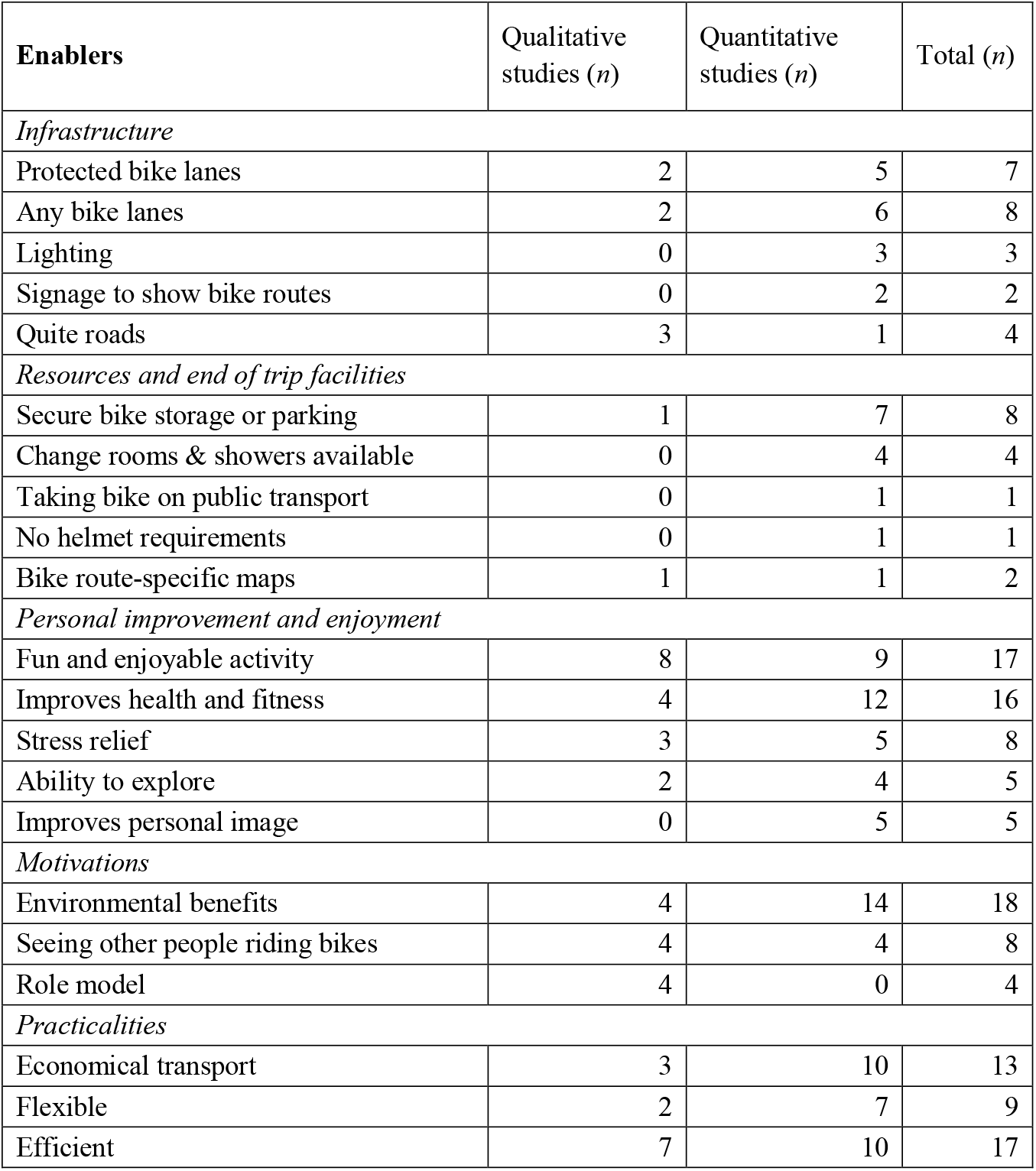
Summary of content analysis in quantitative and qualitative studies and reported enablers

Eight meta-analyses were possible where two or more studies reported unweighted proportional data on the same outcome (Figure 3).

**Figure 3.**
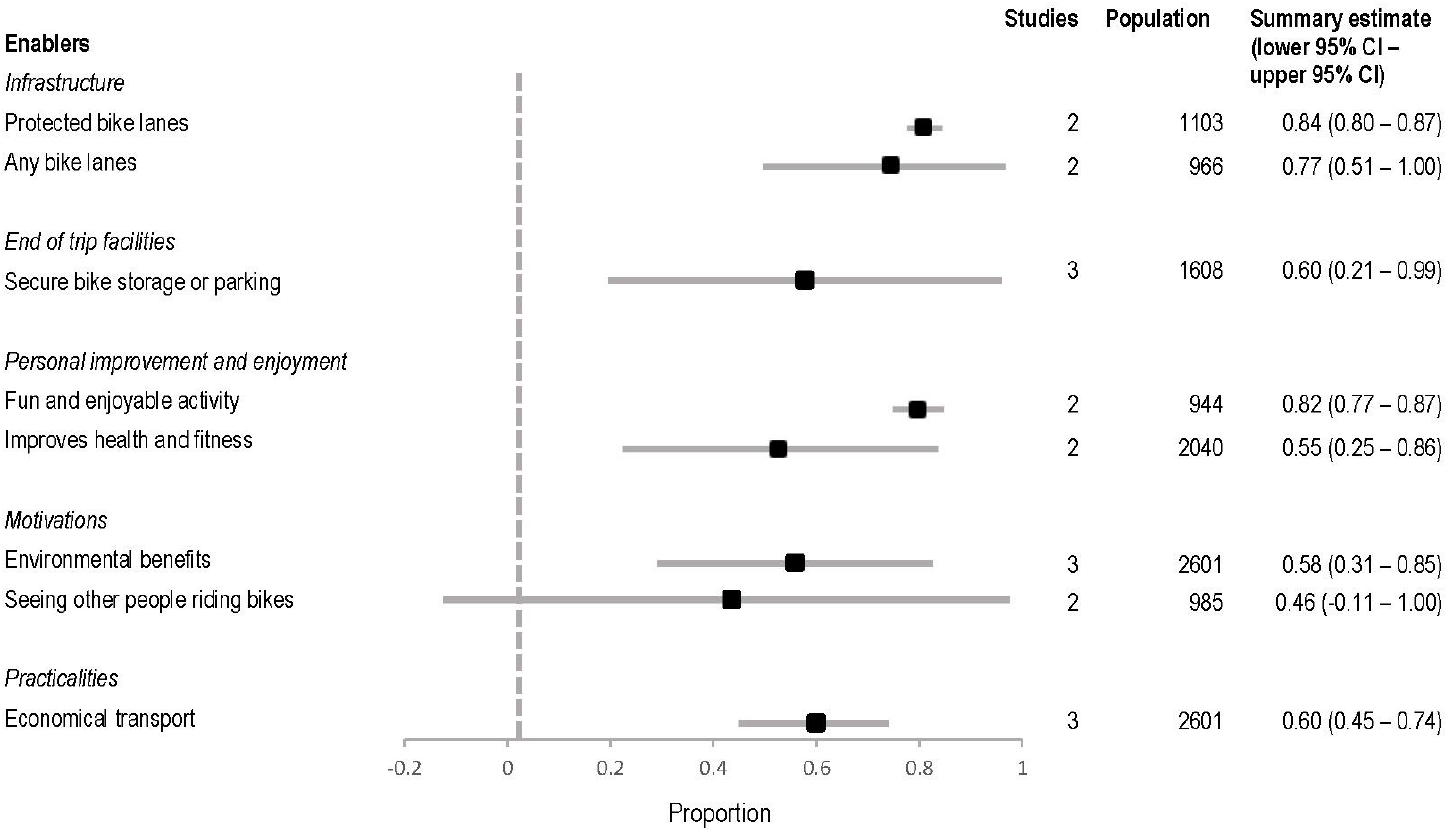
Forest plot of meta-analysis summary estimates (square) and confidence intervals (error bar) of the proportion of people who reported each enabler to cycling for transport

#### Infrastructure

The provision of protected bike lanes (bike lanes physically separated from traffic) had the highest summary estimate relative to other meta-analyses of enablers (summary estimate = 84%; 95% CI: 80%; 87%). This was further rated as important in two Likert scale studies (Crawford et al., 2001; Winters et al., 2011). Adequate lighting, signage specific to people riding bikes, or having a mostly flat route were also rated as important enablers (Crawford et al., 2001; Heart Foundation, 2013; Stangeby, 1997; The Heart Foundation, 2011; Winters et al., 2011).

#### Resources and end of trip facilities

Having access to changing rooms and showers was measured as an enabler in three quantitative studies that were not included in meta-analyses (Crawford et al., 2001; Heart Foundation, 2013; Whannell et al., 2012), where in one survey of women, 4.9% reported this as the main reason that would enable them to ride a bike (Heart Foundation, 2013).

#### Personal improvement and enjoyment

Factors related to personal improvement and enjoyment from bike riding were widely reported in qualitative data. A high summary estimate was reported for riding a bike being a fun and enjoyable activity (summary estimate = 55%; 95% CI: 25%; 86%), further supported by participants across eight studies (Table 6) that commented on the enjoyment of riding a bike.

> “I always feel happy [when e-biking]. I liked it so much better than sitting in a car.” (Jones et al., 2016)

Some studies reported bike riding as a stress release, or to help a person’s mental health, as an enabling factor. In a survey conducted in the centre of Madrid (Muñoz et al., 2013) participants reported a mean of 7.04 on a scale of 1 (strongly disagree) to 10 (strongly agree), for stress release being an enabling factor for bike riding. Participants recalled bike riding as “*my little bit of time alone” (M. Daley et al., 2007)*.

#### Motivations

Bike riding as a more sustainable and environmentally friendly option to travel was reported as less of an enabling factor compared to summary estimates of other enablers (summary estimate = 58%; 95% CI: 31%; 85%). However, this was rated as an important enabling factor across all studies that measured this with a Likert scale (Crawford et al., 2001; De Geus et al., 2008; Fernández-Heredia et al., 2014; Heinen et al., 2011; Muñoz et al., 2013), and one study of women who ride a bike as their primary mode of transport (Cred Consulting, 2020). Participants in four qualitative studies (Galway et al., 2021; Samantha J Leger et al., 2019; D. Lois et al., 2016; J. E. van Bekkum et al., 2011) reported riding a bike because it is *“better for the planet”* (Galway et al., 2021).

An enabling factor reported in four qualitative studies (Rérat, 2019; Salvo et al., 2018; Scott, 2009; J. E. van Bekkum et al., 2011), and not measured in quantitative studies, was being a role model by riding a bike for transport. Participants in several studies commented on riding a bike in hope that their children would do the same in the future.

> “I’d like my son to see that cycling was a viable choice of transport. He’s too ready to jump into the car at every opportunity.” (J. E. van Bekkum et al., 2011)

Potentially related to this enabling factor was the improvement of personal image from riding a bike. This was recorded in five quantitative studies (De Geus et al., 2008; Heinen et al., 2011; Muñoz et al., 2013; Ton et al., 2019), with conflicting findings on the degree of importance.

#### Practicality

Riding a bike being a more economically viable transport option had a high summary estimate relative to other meta-analyses of enablers (summary estimate = 60%; 95% CI: 45%; 74%). Participants in four Likert scale studies rated this as an important enabling factor (Crawford et al., 2001; Fernández-Heredia et al., 2014; Heinen et al., 2011; Muñoz et al., 2013), while participants in three qualitative studies (D. Lois et al., 2016; Scott, 2009; J. E. van Bekkum et al., 2011) reported that bike riding was a one-time cost rather than ongoing costs, such as with a motor vehicle or public transport.

## Discussion

This systematic review and meta-analysis identified a range of barriers and enablers to cycling for transport from 45 studies. Meta-analyses facilitated comparison between barriers and enablers, and the inclusion of qualitative data highlighted potentially under-researched factors and provided a consumer perspective. In total, 34 barriers and 21 enablers were identified. Meta-analyses were possible for 34 of the identified barriers and enablers. Barriers reported by the largest proportions of participants in meta-analyses were related to safety and infrastructure. Furthermore, the enablers reported by the highest proportion of participants were also regarding provision of infrastructure. Barriers identified through qualitative data included a negative perception of bike riders, a negative perception of e-bikes, and a lack of connectivity between existing infrastructure. Qualitative methods identified enablers that were not measured in other quantitative studies, including bike riding permitting exploration of local areas, and riding a bike to act as a role model for children.

Barriers and enablers relating to safety and infrastructure were rated as most important, and reported by the largest number of people in this review. It is likely that barriers regarding a perceived lack of safety relate to the reported lack of infrastructure that supports safe and low-stress bike riding (Branion-Calles et al., 2019). A perceived risk of injury, fear of harassment or aggression from motor vehicle drivers, and fear of riding a bike on the road with cars, were widely reported as barriers to riding a bike. A lack of high-quality infrastructure minimising interactions between bikes and motor vehicles, such as protected bike lanes, were also commonly reported. There is potential for these barriers, which are largely safety and infrastructure-related, to be overcome by interventions that minimise interactions between bike riders and motor vehicle traffic. The provision of high-quality protected infrastructure remains the gold-standard for increasing participation in bike riding (Hull & O’Holleran, 2014; Stewart et al., 2015). However, there is difficulty in providing protected infrastructure across entire cities and regions. Further interventions to minimise potentially unsafe interactions between bike riders and motor vehicles could enable connections between protected infrastructure. This may include areas of lower motor vehicle speeds (such as 30km/h urban speed limits) or traffic calming infrastructure (Yang et al., 2010). Despite the need for protected infrastructure, on-road bike lanes remain a prominent form of bike infrastructure in urban design (Beck, 2021). As supported by our findings where “any bike lane” had a lower summary estimate than specifically protected bike lanes, this type of infrastructure does little to enhance participation in bike riding (Akar & Clifton, 2009; Michelle Daley & Rissel, 2011; J. Dill, 2009; Kristiann C Heesch et al., 2012; Twaddle et al., 2010), and places a person at a substantially higher risk of injury compared to riding a bike in a protected bike lane (Haileyesus et al., 2007; Lusk et al., 2011; Mehan et al., 2009).

A lack of storage on a bike was widely reported as a barrier to riding a bike for transport. This included storage to carry children and objects required for work. This suggests there is a greater need for availability of bikes with enhanced storage capacity, and for environments supportive of such bikes. One way to account for this need could be an increase in the accessibility of ‘cargo bikes’. Cargo bikes have substantial carrying capacity permitting the movement of children and belongings, however there are issues in uptake due to current infrastructure often being too narrow to comfortably ride and manoeuvre in (Liu et al., 2020). Many urban environments are designed with the premise of being used solely by upright two-wheeled bikes (Napper, 2020). Newly established infrastructure dedicated to bikes may benefit from input from cargo-bike riders to ensure lanes are designed to enhance the capacity for bikes with storage capabilities to ride on them.

When designing inclusive infrastructure, it is also important to incorporate the needs of people riding bikes with children (Aldred et al., 2017; Clayton & Musselwhite, 2013). It was evident through qualitative data that some adults would ride a bike for transport if they were able to with their children. However, they reported not feeling safe to do so with existing infrastructure. Other research indicates that parents often avoid riding a bike for travel as sections of their bike network are perceived as unsafe for children, such as sections with on-road lanes (McLaren, 2016). While the barriers to children riding a bike to commute to school have been thoroughly researched, there is limited data on the barriers to adults riding alongside children for other trips. As many trips with children are often within local areas (Yeung et al., 2008), there is potential for modal shifts to bike riding if the needs of this group are understood and met.

A range of measures were used in studies included in this review to identify barriers or enablers, or quantify the importance of these barriers and enablers. Because of this, many study findings were unable to be robustly compared or synthesised, limiting discussion of differences in barriers and enablers between countries, samples and study types. Measures included in meta-analyses used binary outcomes which were useful for showing the size of the effect of a particular barrier or enabler. However, this does not consider the relative strength that a barrier or enabler may have in influencing an individual’s bike riding behaviours. Other studies used Likert scales, permitting the participant to choose if a factor was very much, or less so of a barrier or enabler to their riding a bike. This approach highlights potentially influential factors, allowing for a more focused approach. A downside of using Likert scales in this context is that studies exploring barriers and enablers to bike riding often have a large list of factors for inclusion. Where each factor requires a Likert scale answer, this can contribute to respondent fatigue, threatening data validity (Cox III, 1980; Dolnicar et al., 2011). To allow for comparisons between, and synthesis of, study findings in future, an approach that utilises the benefits of both binary and Likert scale measures is necessary. A two-step logic where participants first select all factors that are a barrier or enabler to them riding a bike, followed by a Likert scale question displaying only those factors chosen by the participant could be beneficial. This approach would identify both the proportion of people affected by particular barriers and enablers, and how much they do so.

It was evident through qualitative findings that there were varying interpretations of questions regarding feelings of safety while riding a bike. Some people perceived questions around feelings of safety to mean feeling unsafe riding a bike, while others perceived this as not feeling a sense of personal safety in, for example, a particular area. The separation of safety due to traffic and road safety compared to safety due to crime, theft or other environmental factors in future research is imperative for the validity and reliability of data (Forman et al., 2008). Pikora et al. (2002) previously outlined potential questions and keywords to specify and reliably measure safety in bike riding research, however this could benefit from updated review (Pikora et al., 2002).

Some enablers reported in this review were identified exclusively through qualitative data. These included riding a bike for transport as a role model for children, in a hope that they will see it as a viable choice of transport. This enabling factor is also present in qualitative research of the enablers of physical activity (Caperchione et al., 2012; Stronach et al., 2016). As it is well established that parents participating in physical activity will have a positive influence on children’s perception and participation in physical activity (Cheung & Chow, 2010; Welk et al., 2003), and there is evidence that this may be an enabling factor for adults, further quantitative measurement and exploration may be beneficial. An understanding of enabling factors such as this have the potential to inform meaningful promotional campaigns and values-based messaging around bike riding.

Reviews in bike riding and active transport research are often limited to published research, excluding potentially relevant grey literature conducted as part of government or bicycle organisation research. This review included a systematic grey literature search to incorporate this, while maintaining rigour by using study-specific quality assessment tools. While consideration was taken to carefully interpret data from grey literature sources, as they were not peer-reviewed, these sources may be subject to selection and publication bias.

A further limitation extends from the synthesis of data across a diversity of populations and locations. Locations had varying geographical areas, trip distances and infrastructure. Populations varied, including groups of university students, and studies of solely women. These variations and the substantial heterogeneity between samples prevented robust comparisons being made between study designs, countries and populations. Despite these variations and their likely influence on results, certain summary estimates had smaller confidence intervals relative to others. This demonstrates the consistency of reporting these barriers and enablers across diverse geographical areas and population groups. In addition, not all barriers and enablers were able to be meta-analysed due to the heterogeneity of outcome measures used.

This review only included studies conducted in OECD countries and published in the English language. Findings are likely not reflective of the barriers and enablers present for people in lower to middle income economies (Bank, 2019), who encounter vastly different road and infrastructure conditions (Forjuoh, 2003). Further review is required to understand the unique barriers and enablers faced by these areas.

There are other methods used to capture barriers and enablers to cycling than what has been included in this review. Stated preference surveys have been used and reviewed previously, however the majority report on focused categories of barriers and/or enablers, such as infrastructure types (Aldred et al., 2017) or bike parking (Heinen & Buehler, 2019). This review focused on self-reported barriers and enablers from survey and qualitative data, which enabled the capture of the diversity and breadth of barriers and enablers people experience for riding a bike, as well as comparisons in frequency and distribution between categories.

## Conclusion

Understanding the barriers and enablers to cycling for transport is integral to designing environments supportive of current and potential bike riders, for promotional campaigns to encourage participation in bike riding and to influence the development of supportive policy. This systematic review identified a diverse range of barriers and enablers using quantitative and qualitative data, highlighting the importance of safety and infrastructure, and potential flaws in survey reliability. Further research should include measures to show the strength of particular barriers and enablers. It is recommended that future research of barriers to bike riding clearly specifies the meaning of “safety”.

## Data Availability

All data produced in the present study are available upon reasonable request to the authors

