## Supplementary Materials for "Adults’ self-reported barriers and enablers to riding a bike for transport: a systematic review"

### Supplementary Materials A – Quality appraisal of studies

*Table 1. Quality appraisal of studies included and excluded*

| First author, year | Tool used | Total appraisal score | Exclusion reasoning |
|---|---|---|---|
| Abasahl et al. (2018) | JBI Cross-sectional | 57% | |
| Agarwal & North (2012) | JBI Cross-sectional | 100% | |
| Akar et al. (2013) | JBI Cross-sectional | 71% | |
| Biernat (2018) | JBI Cross-sectional | 33% | Causation implied from cross-sectional design |
| Chowdhury & Costello (2016) | JBI Cross-sectional | 66% | |
| Central Goldfields Shire Council (2017) | AACODS | 80% | |
| Crawford et al. (2001) | JBI Cross-sectional | 71% | |
| Cred Consulting (2020) | AACODS | 80% | Unclear reporting of participant numbers |
| Curto et al. (2016) | JBI Cross-sectional | 86% | |
| Daley et al. (2007) | JBI Qualitative | 70% | |
| de Cunha & da Silva (2018) | JBI Cross-sectional | 86% | |
| de Geus et al. (2008) | JBI Cross-sectional | 83% | |
| Delso et al. (2018) | JBI Cross-sectional | 66% | |
| Dill & McNeil (2016) | JBI Cross-sectional | 83% | |
| Fernández-Heredia et al. (2014) | JBI Cross-sectional | 66% | |
| Galway et al. (2021) | JBI Qualitative | 80% | |
| Heart Foundation on behalf of the Cycling Promotion Fund (2011) | AACODS | 70% | |
| Heart Foundation on behalf of the Cycling Promotion Fund (2013) | AACODS | 70% | |
| Heesch et al. (2012) | JBI Cross-sectional | 75% | |

| | | | |
|---|---|---|---|
| Heinen et al. (2011) | JBI Cross-sectional | 63% | |
| Ingunn Stangeby on behalf of Transportøkonomisk institutt (1997) | AACODS | 70% | |
| Iwinska et al. (2018) | JBI Cross-sectional | 82% | |
| Jackson (1998) | AACODS | 40% | Limited description of recruitment, participant demographics and outcome measures |
| Jones et al. (2016) | JBI Qualitative | 70% | |
| Kaplan (2013) | JBI Cross-sectional | 38% | Limited description of recruitment and participant demographics |
| Kelarestaghi (2019) | JBI Cross-sectional | 38% | Limited description of recruitment and participant demographics |
| Lechaud (2016) | AACODS | 70% | |
| Leger et al. (2019) | JBI Qualitative | 90% | |
| Lois et al. (2016) | JBI Qualitative | 90% | |
| Manaugh et al. (2016) | JBI Cross-sectional | 75% | |
| McManus et al. (2005) | JBI Cross-sectional | 63% | |
| Moritz (1997) | JBI Cross-sectional | 38% | Limited description of recruitment and participant demographics |
| Mullan (2013) | JBI Qualitative | 80% | |
| Mullan (2012) | JBI Cross-sectional | 40% | Limited description of recruitment, participant demographics and qualitative analysis |
| Munoz et al. (2013) | JBI Cross-sectional | 63% | |
| Nikitas (2018) | JBI Cross-sectional | 75% | |
| Ransdell (2013) | JBI Cross-sectional | 40% | Limited description of recruitment, participant demographics and outcome measures |
| Rérat (2019) | JBI Cross-sectional | 82% | |
| Sahlqvist & Heesch (2012) | JBI Cross-sectional | 75% | |
| Salvo et al. (2018) | JBI Cross-sectional | 77% | |

| | | | |
|---|---|---|---|
| Schneider & Hu (2015) | JBI Cross-sectional | 87% | |
| Scott (2009) | AACODS | 80% | |
| Sears et al. (2012) | JBI Cohort | 77% | |
| Štastná et al. (2018) | JBI Cross-sectional | 75% | |
| Swiers et al. (2017) | JBI Cross-sectional | 80% | |
| Ton et al. (2019) | JBI Cross-sectional | 88% | |
| Unwin (1992) | JBI Cross-sectional | 38% | Limited description of recruitment, participant demographics and outcome measures |
| van Bekkum et al. (2011) | JBI Cross-sectional | 88% | |
| van Bekkum et al. (2011) | JBI Qualitative | 80% | |
| Webster (2013) | AACODS | 48% | Limited description of participations and limited information on reporting of qualitative results |
| Whannell et al. (2011) | JBI Cross-sectional | 62% | |
| Winters et al. (2011) | JBI Cross-sectional | 100% | |
| Winters et al. (2015) | JBI Qualitative | 100% | |
| Zander et al. (2013) | JBI Qualitative | 80% | |

AACODS: “Authority, Accuracy, Coverage, Objectivity, Date and Significance” Checklist

JBI: Joanna Briggs Institute Quality Appraisal Checklists

Supplementary Materials B - Quantitative data of barriers to cycling for transport

Note: Qualitative data of barriers and enablers can be made available to the reader on request to the corresponding author.

*Table 2. Quantitative data for safety barriers to cycling for transport*

| **First Author, Year** | **Population** | **Measure** | **Perceived risk of injury** | **Feelings of safety** | **Fear of motorist aggression** | **High traffic density** | **Fear of theft/crime** |
|---|---|---|---|---|---|---|---|
| Fernández-Heredia, 2014 | 76% students, rest staff of a university in Madrid | 6 Point Likert Scale 1 (not important) to 6 (fundamental) | | 6 Point Likert Scale *1 (not important) to 6 (fundamental)* Mean = 4.09 | | | 6 Point Likert Scale *1 (not important) to 6 (fundamental)* Mean = 3.32 |
| Crawford, 2001 | Two workplaces within Glasgow - one university and one hospital | 5 Point Likert Scale *1 (not important) to 5 (very important)* | | | | 5 Point Likert Scale *1 (not important) to 5 (very important)* Mean = 4.1 | |
| Manaugh, 2016 | Staff and students at the McGill University in Montreal. | Proportions AND Likert (strongly disagree = 1 to strongly agree = 5) - only from potential cyclists | | 5 Point Likert Scale *1 (strongly disagree) to 5 (strongly agree)* Mean = 3.1 | | | |
| Rérat, 2019 | People who participated in the Bike to Work campaign in Switzerland | Proportion that reported "strongly agree" or 'slightly agree' for factor being a barrier | Proportion that reported "strongly agree" or "slightly agree" n = 36% | | | | Proportion that reported "strongly agree" or "slightly agree" n = 26% |
| Swiers, 2018 | Students at an urban UK university | Proportion that reported "strongly agree" or 'slightly agree' for factor being a barrier | | Proportion that reported "strongly agree" or "slightly agree" n = 39% | | | |

| | | | | | | |
|---|---|---|---|---|---|---|
| van Bekkum, 2011 | Staff and PhD students in a cycle-friendly building in a university in Edinburgh. | 5-point Likert scale from 1 = not discouraging to 5 = stops me from cycling (mean, SD) Female, Male data only | | 5 Point Likert Scale by Gender<br>*1 (not discouraging) to 5 (stops me from cycling)*<br><u>Mean (Women) = 3.83</u><br><u>Mean (Men) = 3.18</u> | | |
| Winters, 2011 | Current and potential cyclists in Vancouver from a random sample of home addresses | Likert scale from -1 (much less likely to cycle) to 1 (much more likely to cycle) | 3 Point Likert Scale<br>*-1 (much less likely to cycle) to 2 (much more likely to cycle)*<br><u>Mean = -0.67</u> | | 3 Point Likert Scale<br>*-1 (much less likely to cycle) to 2 (much more likely to cycle)*<br><u>Mean = -0.73</u> | 3 Point Likert Scale<br>*-1 (much less likely to cycle) to 2 (much more likely to cycle)*<br><u>Mean = -0.83</u> |
| Scott, 2009 | People living in NSW - here only infrequent/non-cyclists who participated in a survey | Weighted data. Proportion of those who selected as reason for not commuting (among those who live within 10km of work/train/ferry) | | Proportion that reported as main reason for not commuting by bike<br><u>n = 23%</u> | | Proportion that reported as main reason for not commuting by bike<br><u>n = 2%</u> |
| The Heart Foundation, 2013 | Adult women mostly residing in urban areas | Reasons that prevent women from cycling - proportion that reported as main reason | Proportion that reported as main reason for not commuting by bike<br><u>n = 5.8%</u> | | Proportion that reported as main reason for not commuting by bike<br><u>n = 7.1%</u> | Proportion that reported as main reason for not commuting by bike<br><u>n = 8.2%</u> |
| Lechaud, 2016 | Convenience sample of 701 people living in Portugal | Proportion that selected slightly or completely agree for main motives to not commute by bike | | Proportion that reported "completely agree" or "slightly agree"<br><u>n = 44%</u> | | |

| | | | | | | |
|---|---|---|---|---|---|---|
| Dill, 2016 | People living in the USA who had not ridden a bike in the last 30 days (n = 2563) | Proportion that selected barrier to bicycling more | | Proportion that reported as barrier to not bicycling more<br>Strong and fearless group = 12%<br>Enthused and confident group = 18%<br>Interested but concerned = 15% | | Proportion that reported as barrier to not bicycling more<br>Strong and fearless group = 26%<br>Enthused and confident group = 31%<br>Interested but concerned = 46% |
| Delso, 2018 | Short car trip user who could be a potential cyclist or walker (SCT), Short car trip users & bicycle users, bicycle users | Likert scale 1-7 (1 not limiting to 7 very limiting) | | 7 Point Likert Scale<br>1 (not limiting) to 7 (very limiting)<br>SCT Mean = 4.92<br>SCT & Bicycle User Mean = 4.65<br>Bicycle User Mean = 3.88 | | 7 Point Likert Scale<br>1 (not limiting) to 7 (very limiting)<br>SCT Mean = 6.17<br>SCT & Bicycle User Mean = 5.89<br>Bicycle User Mean = 4.88 |

*Table 3. Quantitative data for infrastructure barriers to cycling for transport*

| **Author, Year** | **Population** | **Measure** | **Limited dedicated bike lanes** | **Poor quality and condition of dedicated bike lanes** | **Poor condition of roads** | **Darkness** |
|---|---|---|---|---|---|---|
| Crawford, 2001 | Two workplaces within Glasgow - one university and one hospital | 5 Point Likert Scale<br>*1 (not important) to 5 (very important)* | | | 5 Point Likert Scale<br>*1 (not important) to 5 (very important)*<br>Mean = 3.7 | |
| Sears, 2012 | Group of working adults in Vermont who commute by bicycle two or more miles each way on 28 specified days in a 10-month period | Percentage of commuting days not biked for this reason (frequency cited - person days) | | | | Proportion of commuting days not biked for this reason<br>n = 11.6% |
| van Bekkum, 2011 | Staff and PhD students in a cycle-friendly building in a university in Edinburgh. | 5-point Likert scale from 1 = not discouraging to 5 = stops me from cycling | | | 5 Point Likert Scale by Gender<br>*1 (not discouraging) to 5 (stops me from cycling)* | 5 Point Likert Scale by Gender<br>*1 (not discouraging) to 5 (stops me from cycling)* |

| | | | | | | |
|---|---|---|---|---|---|---|
| | | (mean, SD) Female, Male data only | | | Mean (Women) = 2.54<br>Mean (Men) = 2.14 | Mean (Women) = 2.90<br>Mean (Men) = 2.24 |
| Winters, 2011 | Current and potential cyclists in Vancouver from a random sample of home addresses | Likert scale from -1 (much less likely to cycle) to 1 (much more likely to cycle) | | | 3 Point Likert Scale<br>*-1 (much less likely to cycle) to 2 (much more likely to cycle)*<br>Mean = -0.76 | 3 Point Likert Scale<br>*-1 (much less likely to cycle) to 2 (much more likely to cycle)*<br>Mean = -0.59 |
| Scott, 2009 | People living in NSW - here only infrequent/non-cyclists who participated in a survey | Weighted data. Proportion of those who selected as reason for not commuting (among those who live within 10km of work/train/ferry) | Proportion that reported as main reason for not commuting by bike<br>n = 3% | | | |
| The Heart Foundation, 2013 | Adult women mostly residing in urban areas | Reasons that prevent women from cycling - proportion that reported as main reason | Proportion that reported as main reason for not commuting by bike<br>n = 7.1% | | | |
| Lechaud, 2016 | Convenience sample of 701 people living in Portugal | Proportion that selected slightly or completely agree for main motives to not commute by bike | Proportion that reported "completely agree" or "slightly agree"<br>n = 55% | Proportion that reported "completely agree" or "slightly agree"<br>n = 55% | | |
| Dill, 2016 | People living in the USA who had not ridden a bike in the last 30 days (n = 2563) | Proportion that selected barrier to bicycling more | Proportion that reported as barrier to not bicycling more<br>Strong and fearless group = 22%<br>Enthused and confident group = 44%<br>Interested but concerned = 43% | | | |
| Delso, 2018 | Short car trip user who could be a potential cyclist or walker (SCT), Short car trip users & bicycle users, bicycle users | Likert scale 1-7 (1 not limiting to 7 very limiting) | 7 Point Likert Scale<br>1 (not limiting) to 7 (very limiting)<br>SCT Mean = 5.14<br>SCT & Bicycle User Mean = 4.7<br>Bicycle User Mean = 4.39 | | | |

*Table 4. Quantitative data for trip factors barriers to cycling for transport*

| **Author, Year** | **Population** | **Measure used** | **Long distance** | **Time to destination too great** | **Other activities before/after work** | **Lack of storage on bike** | **Need vehicle for work** | **Children to transport** |
|---|---|---|---|---|---|---|---|---|
| de Geus, 2007 | Working Flemish adults who lived within 10km of their workplace | Five-point scale from 1 (strongly disagree) to 5 (strongly agree) | | 5 Point Likert Scale<br>*1 (strongly disagree) - 5 (strongly agree)*<br><u>Mean = 2.6</u> | | | | |
| Fernández-Heredia, 2014 | 76% students, rest staff of a university in Madrid | 6 Point Likert Scale<br>1 (not important) to 6 (fundamental) | 6 Point Likert Scale<br>*1 (not important) to 6 (fundamental)*<br><u>Mean = 3.61</u> | | | | | |
| Crawford, 2001 | Two workplaces within Glasgow - one university and one hospital | 5 Point Likert Scale<br>*1 (not important) to 5 (very important)* | 5 Point Likert Scale<br>*1 (not important) to 5 (very important)*<br><u>Mean = 3.8</u> | 5 Point Likert Scale<br>*1 (not important) to 5 (very important)*<br><u>Mean = 3.8</u> | | | | 5 Point Likert Scale<br>*1 (not important) to 5 (very important)*<br><u>Mean = 3.7</u> |
| Manaugh, 2016 | Staff and students at the McGill University in Montreal. | Proportions AND Likert (strongly disagree = 1 to strongly agree = 5) - only from potential cyclists | 5 Point Likert Scale<br>*1 (strongly disagree) to 5 (strongly agree)*<br><u>Mean = 2.3</u> | | | | | |
| Rérat, 2019 | People who participated in the Bike to Work campaign in Switzerland | Proportion that reported "strongly agree" or 'slightly agree' | | Proportion that reported "strongly agree" or "slightly agree"<br><u>n = 19%</u> | Proportion that reported "strongly agree" or "slightly agree"<br><u>n = 40%</u> | Proportion that reported "strongly agree" or "slightly agree"<br><u>n = 47%</u> | | Proportion that reported "strongly agree" or "slightly agree"<br><u>n = 21%</u> |

| | | | | | | | | |
|---|---|---|---|---|---|---|---|---|
| Sears, 2012 | Group of working adults in Vermont who commute by bicycle two or more miles each way on 28 specified days in a 10-month period | Percentage of commuting days not biked for this reason (frequency cited - person days) | | | Proportion of commuting days not biked for this reason<br>n = 16.5% | Proportion of commuting days not biked for this reason<br>n = 4.6% | | Proportion of commuting days not biked for this reason<br>n = 4.8% |
| van Bekkum, 2011 | Staff and PhD students in a cycle-friendly building in a university in Edinburgh. | 5-point Likert scale from 1 = not discouraging to 5 = stops me from cycling (mean, SD) Female, Male data only | 5 Point Likert Scale by Gender<br>*1 (not discouraging) to 5 (stops me from cycling)*<br>Mean (Women) = 2.34<br>Mean (Men) = 2.06 | 5 Point Likert Scale by Gender<br>*1 (not discouraging) to 5 (stops me from cycling)*<br>Mean (Women) = 2.08<br>Mean (Men) = 1.90 | | 5 Point Likert Scale by Gender<br>*1 (not discouraging) to 5 (stops me from cycling)*<br>Mean (Women) = 2.29<br>Mean (Men) = 1.89 | | 5 Point Likert Scale by Gender<br>*1 (not discouraging) to 5 (stops me from cycling)*<br>Mean (Women) = 2.48<br>Mean (Men) = 1.66 |
| Winters, 2011 | Current and potential cyclists in Vancouver from a random sample of home addresses | Likert scale from -1 (much less likely to cycle) to 1 (much more likely to cycle) | | | | 3 Point Likert Scale<br>*-1 (much less likely to cycle) to 2 (much more likely to cycle)*<br>Mean = -0.57 | | |
| Scott, 2009 | People living in NSW - here only infrequent/non-cyclists who participated in a survey | Weighted data. Proportion of those who selected as reason for not commuting (among those who live within 10km of work/train/ferry) | | Proportion that reported as main reason for not commuting by bike<br>n = 5% | | | Proportion that reported as main reason for not commuting by bike<br>n = 1% | |
| The Heart Foundation, 2013 | Adult women mostly residing in urban areas | Reasons that prevent women from cycling - proportion that reported as main reason | Proportion that reported as main reason for not commuting by bike<br>n = 5.9% | Proportion that reported as main reason for not commuting by bike<br>n = 10% | | Proportion that reported as main reason for not commuting by bike<br>n = 4% | | Proportion that reported as main reason for not commuting by bike<br>n = 6.3% |

| | | | | | | | |
|---|---|---|---|---|---|---|---|
| Dill, 2016 | People living in the USA who had not ridden a bike in the last 30 days (n = 2563) | Proportion that selected barrier to bicycling more | Proportion that reported as barrier to not bicycling more<br>Strong and fearless group = 36%<br>Enthused and confident group = 45%<br>Interested but concerned = 50% | | | | Proportion that reported as barrier to not bicycling more<br>Strong and fearless group = 54%<br>Enthused and confident group = 63%<br>Interested but concerned = 53% |
| Delso, 2018 | Short car trip user who could be a potential cyclist or walker (SCT), Short car trip users & bicycle users, bicycle users | Likert scale 1-7 (1 not limiting to 7 very limiting) | 7 Point Likert Scale<br>1 (not limiting) to 7 (very limiting)<br>SCT Mean = 5.16<br>SCT & Bicycle User Mean = 4.87<br>Bicycle User Mean = 3.83 | | | | |

*Table 5. Quantitative data for personal barriers to cycling for transport*

| | | | | | | | | |
|---|---|---|---|---|---|---|---|---|
| de Geus, 2007 | Working Flemish adults who lived within 10km of their workplace | Five-point scale from 1 (strongly disagree) to 5 (strongly agree) | | | | | 5 Point Likert Scale<br>*1 (strongly disagree) - 5 (strongly agree)*<br>Mean 2.58 | |
| Fernández-Heredia, 2014 | 76% students, rest staff of a university in Madrid | 6 Point Likert Scale<br>1 (not important) to 6 (fundamental) | | | | 6 Point Likert Scale<br>*1 (not important) to 6 (fundamental)*<br>Mean = 2.46 | | 6 Point Likert Scale<br>*1 (not important) to 6 (fundamental)*<br>Mean = 3.18 |

| | | | | | | | | | | |
|---|---|---|---|---|---|---|---|---|---|---|
| Crawford, 2001 | Two workplaces within Glasgow - one university and one hospital | 5 Point Likert Scale *1 (not important) to 5 (very important)* | | | | 5 Point Likert Scale *1 (not important) to 5 (very important)* Mean = 2.6 | | | | 5 Point Likert Scale *1 (not important) to 5 (very important)* Mean = 2.6 |
| Manaugh, 2016 | Staff and students at the McGill University in Montreal. | Proportions AND Likert (strongly disagree = 1 to strongly agree = 5) - only from potential cyclists | | | 5 Point Likert Scale *1 (strongly disagree) to 5 (strongly agree)* Mean = 2.9 | | | 5 Point Likert Scale *1 (strongly disagree) to 5 (strongly agree)* Mean = 2.5 | | |
| Rérat, 2019 | People who participated in the Bike to Work campaign in Switzerland | Proportion that reported "strongly agree" or 'slightly agree' | | Proportion that reported "strongly agree" or "slightly agree" n = 19% | Proportion that reported "strongly agree" or "slightly agree" n = 16% | | | | | Proportion that reported "strongly agree" or "slightly agree" n = 20% |
| Sears, 2012 | Group of working adults in Vermont who commute by bicycle two or more miles each way on 28 specified days in a 10-month period | Percentage of commuting days not biked for this reason (frequency cited - person days) | | | Proportion of commuting days not biked for this reason n = 2.9% | | | | | Proportion of commuting days not biked for this reason n = 0.7% |
| van Bekkum, 2011 | Staff and PhD students in a cycle-friendly building in a university in Edinburgh. | 5-point Likert scale from 1 = not discouraging to 5 = stops me from cycling (mean, SD) | | 5 Point Likert Scale by Gender *1 (not discouraging) to 5 (stops me from cycling)* Mean (Women) = | 5 Point Likert Scale by Gender *1 (not discouraging) to 5 (stops me from cycling)* Mean (Women) = | | | | 5 Point Likert Scale by Gender *1 (not discouraging) to 5 (stops me from cycling)* Mean (Women) = | |

| | | | | | | | | | |
|---|---|---|---|---|---|---|---|---|---|
| | | Female, Male data only | | 1.84<br>Mean (Men) = 1.41 | 1.96<br>Mean (Men) = 1.55 | | | | 1.59<br>Mean (Men) = 1.44 |
| Scott, 2009 | People living in NSW - here only infrequent/non-cyclists who participated in a survey | Weighted data. Proportion of those who selected as reason for not commuting | Proportion that reported as main reason for not commuting by bike<br>n = 1% | Proportion that reported as main reason for not commuting by bike<br>n = 5% | Proportion that reported as main reason for not commuting by bike<br>n = 3% | | Proportion that reported as main reason for not commuting by bike<br>n = 2% | | Proportion that reported as main reason for not commuting by bike<br>n = 3% |
| The Heart Foundation, 2013 | Adult women mostly residing in urban areas | Reasons that prevent women from cycling - proportion that reported as main reason | Proportion that reported as main reason for not commuting by bike<br>n = 0.4% | Proportion that reported as main reason for not commuting by bike<br>n = 7.6% | | | | | |
| Lechaud, 2016 | Convenience sample of 701 people living in Portugal | Proportion that selected slightly or completely agree for main motives to not commute by bike | | | | | | Proportion that reported "completely agree" or "slightly agree"<br>n = 15% | |
| Delso, 2018 | Short car trip user who could be a potential cyclist or walker (SCT), Short car trip users & bicycle users, bicycle users | Likert scale 1-7 (1 not limiting to 7 very limiting) | 7 Point Likert Scale<br>1 (not limiting) to 7 (very limiting)<br>SCT Mean = 5.71<br>SCT & Bicycle User Mean = 5.17<br>Bicycle User Mean = 3.91 | | | 7 Point Likert Scale<br>1 (not limiting) to 7 (very limiting)<br>SCT Mean = 4.47<br>SCT & Bicycle User Mean = 3.47<br>Bicycle User Mean = 2.86 | | | |

*Table 6.Quantitative data for access barriers to cycling for transport*

| **Author, Year** | **Population** | **Measure used** | **No bike** | **Cost of bike too high** | **Mandatory helmet laws** |
|---|---|---|---|---|---|
| Crawford, 2001 | Two workplaces within Glasgow - one university and one hospital | 5 Point Likert Scale<br>*1 (not important) to 5 (very important)* | | 5 Point Likert Scale<br>*1 (not important) to 5 (very important)*<br><u>Mean = 3.7</u> | |
| Manaugh, 2016 | Staff and students at the McGill University in Montreal. | Proportions AND Likert (strongly disagree = 1 to strongly agree = 5) - only from potential cyclists | | 5 Point Likert Scale<br>*1 (strongly disagree) to 5 (strongly agree)*<br><u>Mean = 1.6</u> | |
| van Bekkum, 2011 | Staff and PhD students in a cycle-friendly building in a university in Edinburgh. | 5-point Likert scale from 1 = not discouraging to 5 = stops me from cycling (mean, SD) Female, Male data only | | 5 Point Likert Scale by Gender<br>*1 (not discouraging) to 5 (stops me from cycling)*<br><u>Mean (Women) = 1.85</u><br><u>Mean (Men) = 1.61</u> | |
| Scott, 2009 | People living in NSW - here only infrequent/non-cyclists who participated in a survey | Weighted data. Proportion of those who selected as reason for not commuting (among those who live within 10km of work/train/ferry) | | Proportion that reported as main reason for not commuting by bike<br><u>n = 7%</u> | |
| Dill, 2016 | People living in the USA who had not ridden a bike in the last 30 days (n = 2563) | Proportion that selected barrier to bicycling more | Proportion that reported as barrier to not bicycling more<br>Strong and fearless group = 46%<br>Enthused and confident group = 51%<br>Interested but concerned = 56% | | |
| Delso, 2018 | Short car trip user who could be a potential cyclist or walker (SCT), Short car trip users & bicycle users, bicycle users | Likert scale 1-7 (1 not limiting to 7 very limiting) | | | 7 Point Likert Scale<br>1 (not limiting) to 7 (very limiting)<br>SCT Mean = 3.34<br>SCT & Bicycle User Mean = 3.45<br>Bicycle User Mean = 3.57 |

*Table 7. Quantitative data for environmental barriers to cycling for transport*

| Author, Year | Population | Measure used | Air-pollution exposure | Bad weather | Too many hills |
|---|---|---|---|---|---|
| Fernández-Heredia, 2014 | 76% students, rest staff of a university in Madrid | 6 Point Likert Scale 1 (not important) to 6 (fundamental) | | 6 Point Likert Scale *1 (not important) to 6 (fundamental)* Mean = 3.63 | 6 Point Likert Scale *1 (not important) to 6 (fundamental)* Mean = 3.42 |
| Crawford, 2001 | Two workplaces within Glasgow - one university and one hospital | 5 Point Likert Scale *1 (not important) to 5 (very important)* | 5 Point Likert Scale *1 (not important) to 5 (very important)* Mean = 3.8 | 5 Point Likert Scale *1 (not important) to 5 (very important)* Mean = 4.3 | 5 Point Likert Scale *1 (not important) to 5 (very important)* Mean = 3.7 |
| Rérat, 2019 | People who participated in the Bike to Work campaign in Switzerland | Proportion that reported "strongly agree" or 'slightly agree' | Proportion that reported "strongly agree" or "slightly agree" n = 24% | Proportion that reported "strongly agree" or "slightly agree" n = 53% | |
| Sears, 2012 | Group of working adults in Vermont who commute by bicycle two or more miles each way on 28 specified days in a 10-month period | Percentage of commuting days not biked for this reason (frequency cited - person days) | | Proportion of commuting days not biked for this reason n = 16.5% | |
| Swiers, 2018 | Students at an urban UK university | Proportion that reported "strongly agree" or "slightly agree" as preventing them from riding a bike | | Proportion that reported "strongly agree" or "slightly agree" n = 62% | |
| van Bekkum, 2011 | Staff and PhD students in a cycle-friendly building in a university in Edinburgh. | 5-point Likert scale from 1 = not discouraging to 5 = stops me from cycling (mean, SD) Female, Male data only | 5 Point Likert Scale by Gender *1 (not discouraging) to 5 (stops me from cycling)* Mean (Women) = 2.56 Mean (Men) = 2.10 | 5 Point Likert Scale by Gender *1 (not discouraging) to 5 (stops me from cycling)* Mean (Women) = 3.21 Mean (Men) = 2.8 | 5 Point Likert Scale by Gender *1 (not discouraging) to 5 (stops me from cycling)* Mean (Women) = 2.69 Mean (Men) = 2 |
| Winters, 2011 | Current and potential cyclists in Vancouver from a random sample of home addresses | Likert scale from -1 (much less likely to cycle) to 1 (much more likely to cycle) | | 3 Point Likert Scale *-1 (much less likely to cycle) to 2 (much more likely to cycle)* Mean = -0.63 | |
| Scott, 2009 | People living in NSW - here only infrequent/non-cyclists who participated in a survey | Weighted data. Proportion of those who selected as reason for not commuting (among those who live within 10im of work/train/ferry) | Proportion that reported as main reason for not commuting by bike n = 1% | Proportion that reported as main reason for not commuting by bike n = 6% | |

| | | | | | |
|---|---|---|---|---|---|
| The Heart Foundation, 2013 | Adult women mostly residing in urban areas | Reasons that prevent women from cycling - proportion that reported as main reason | Proportion that reported as main reason for not commuting by bike<br>n = 2.8% | Proportion that reported as main reason for not commuting by bike<br>n = 3.1% | |
| Lechaud, 2016 | Convenience sample of 701 people living in Portugal | Proportion that selected slightly or completely agree for main motives to not commute by bike | | Proportion that reported "completely agree" or "slightly agree"<br>n = 27% | Proportion that reported "completely agree" or "slightly agree"<br>n = 38% |
| Dill, 2016 | People living in the USA who had not ridden a bike in the last 30 days (n = 2563) | Proportion that selected barrier to bicycling more | | Proportion that reported as barrier to not bicycling more<br>Strong and fearless group = 28%<br>Enthused and confident group = 36%<br>Interested but concerned = 38% | |
| Delso, 2018 | Short car trip user who could be a potential cyclist or walker (SCT), Short car trip users & bicycle users, bicycle users | Likert scale 1-7 (1 not limiting to 7 very limiting) | | | 7 Point Likert Scale<br>1 (not limiting) to 7 (very limiting)<br>SCT Mean = 5.08<br>SCT & Bicycle User Mean = 4.37<br>Bicycle User Mean = 3.45 |

*Table 8. Quantitative data for end of trip barriers to cycling for transport*

| **Author, Year** | **Population** | **Measure used** | **Lack of bike parking** | **Lack of showers or lockers** |
|---|---|---|---|---|
| Fernández-Heredia, 2014 | 76% students, rest staff of a university in Madrid | 6 Point Likert Scale<br>1 (not important) to 6 (fundamental) | 6 Point Likert Scale<br>*1 (not important) to 6 (fundamental)*<br>Mean = 4.43 | |
| Crawford, 2001 | Two workplaces within Glasgow - one university and one hospital | 5 Point Likert Scale<br>*1 (not important) to 5 (very important)* | | 5 Point Likert Scale<br>*1 (not important) to 5 (very important)*<br>Mean = 3.7 |
| Manaugh, 2016 | Staff and students at the McGill University in Montreal. | Proportions AND Likert (strongly disagree = 1 to strongly agree = 5) - only from potential cyclists | 5 Point Likert Scale<br>*1 (strongly disagree) to 5 (strongly agree)*<br>Mean = 3.1 | |

| | | | | |
|---|---|---|---|---|
| Swiers, 2018 | Students at an urban UK university | | | Proportion that reported "strongly agree" or "slightly agree"<br>n = 24% |
| van Bekkum, 2011 | Staff and PhD students in a cycle-friendly building in a university in Edinburgh. | 5-point Likert scale from 1 = not discouraging to 5 = stops me from cycling (mean, SD) Female, Male data only | 5 Point Likert Scale by Gender<br>*1 (not discouraging) to 5 (stops me from cycling)*<br>Mean (Women) = 1.72<br>Mean (Men) = 1.71 | 5 Point Likert Scale by Gender<br>*1 (not discouraging) to 5 (stops me from cycling)*<br>Mean (Women) = 1.81<br>Mean (Men) = 1.77 |
| Scott, 2009 | People living in NSW - here only infrequent/non-cyclists who participated in a survey | Weighted data. Proportion of those who selected as reason for not commuting (among those who live within 10km of work/train/ferry) | Proportion that reported as main reason for not commuting by bike<br>n = 6% | Proportion that reported as main reason for not commuting by bike<br>n = 6% |
| The Heart Foundation, 2013 | Adult women mostly residing in urban areas | Reasons that prevent women from cycling - proportion that reported as main reason | | Proportion that reported as main reason for not commuting by bike<br>n = 2.3% |

#### Supplementary Materials C – Quantitative data of enablers to cycling for transport

***Table 9. Quantitative data for infrastructure enablers to cycling for transport***

| First author, year | Population | Measure used | Any bike lanes | Protected bike lanes | Connectivity | Quiet roads | Lighting | Signage to show bike routes |
|---|---|---|---|---|---|---|---|---|
| Crawford, 2001 | Two workplaces within Glasgow - one university and one hospital | 5 Point Likert Scale<br>*1 (not important) to 5 (very important)* | 5 Point Likert Scale<br>*1 (not important) to 5 (very important)*<br>Mean = 4.3 | 5 Point Likert Scale<br>*1 (not important) to 5 (very important)*<br>Mean = 4.4 | | | | |
| Cred Consulting, 2020 | Women living in Sydney, Australia who ride a bike for most of their commute | Proportion that reported as a factor that influenced the route they choose to take | Proportion that reported as a factor that influenced the route they choose to take<br>n = 62% | Proportion that reported as a factor that influenced the route they choose to take<br>n = 64% | | Proportion that reported as a factor that influenced the route they choose to take<br>n = 57% | Proportion that reported as a factor that influenced the route they choose to take<br>n = 13% | |
| Whannell, 2011 | First year undergraduate students studying a science, technology and society course focussed on environmental and sustainability issues | Proportion that reported "strongly agree" or "slightly agree" | | Proportion that reported "strongly agree" or "slightly agree"<br>n = 68% | | | | |
| Winters, 2011 | Current and potential cyclists in Vancouver from a random sample of home addresses | 3 Point Likert Scale<br>*-1 (much less likely to cycle) to 1 (much more likely to cycle)* | | 3 Point Likert Scale<br>*-1 (much less likely to cycle) to 1 (much more likely to cycle)*<br>Mean - 0.79 | | | 3 Point Likert Scale<br>*-1 (much less likely to cycle) to 1 (much more likely to cycle)*<br>Mean - 0.5 | 3 Point Likert Scale<br>*-1 (much less likely to cycle) to 1 (much more likely to cycle)*<br>Mean - 0.49 |
| The Heart Foundation, 2013 | Adult women mostly residing in urban areas | Proportion that reported as main reason for encouraging women to cycle | Proportion that reported as main reason for encouraging women to cycle<br>n = 16.2% | Proportion that reported as main reason for encouraging women to cycle<br>n = 32.3% | Proportion that reported as main reason for encouraging women to cycle<br>n = 3.6% | | Proportion that reported as main reason for encouraging women to cycle<br>n = 2.5% | |

| | | | | | | | | |
|---|---|---|---|---|---|---|---|---|
| Transportøkonomisk institutt, 1997 | Mostly middle-aged people with a high level of education and high household income | Proportion that reported as most important improvements to start using a bike | Proportion that reported as most important improvements to start using a bike<br>n = 30% | | | | | Proportion that reported as most important improvements to start using a bike<br>n = 1% |

*Table 10. Quantitative data for resource and end of trip enablers to cycling for transport*

| **First author, year** | **Population** | **Measure used** | **Secure bike storage or parking** | **Changing/ showers available** | **Taking bike on public transport** | **No helmet requirements** | **Bike route-specific maps** |
|---|---|---|---|---|---|---|---|
| Crawford, 2001 | Two workplaces within Glasgow - one university and one hospital | 5 Point Likert Scale<br>*1 (not important) to 5 (very important)* | 5 Point Likert Scale<br>*1 (not important) to 5 (very important)*<br>Mean = 4.1 | 5 Point Likert Scale<br>*1 (not important) to 5 (very important)*<br>Mean = 4 | | | |
| Cred Consulting, 2020 | Women living in Sydney, Australia who ride a bike for most of their commute | Proportion that reported as a factor that influenced the route they choose to take | | Proportion that reported as a factor that influenced the route they choose to take<br>n = 20% | | | |
| Munoz, 2013 | People walking in the centre of Madrid and not tourists. | To what extent to you agree with the following factors in relation to cycling | 10 Point Likert Scale<br>*1 (strongly disagree) to 10 (strongly agree)*<br>Mean = 8.21 | | | | |
| Whannell, 2011 | First year undergraduate students studying a science, technology and society course | Proportion that reported "strongly agree" or "slightly agree" | Proportion that reported "strongly agree" or "slightly agree"<br>n = 76% | Proportion that reported "strongly agree" or "slightly agree"<br>n = 50% | | | |
| Winters, 2011 | Current and potential cyclists in Vancouver from a random sample of home addresses | 3 Point Likert Scale<br>*-1 (much less likely to cycle) to 1 (much more likely to cycle)* | 3 Point Likert Scale<br>*-1 (much less likely to cycle) to 1 (much more likely to cycle)*<br>Mean - 0.49 | | 3 Point Likert Scale<br>*-1 (much less likely to cycle) to 1 (much more likely to cycle)*<br>Mean - 0.5 | | |
| The Heart Foundation, 2013 | Adult women mostly residing in urban areas | Proportion that reported as main reason for encouraging women to cycle | | Proportion that reported as main reason for encouraging women to cycle<br>n = 4.9% | | Proportion that reported as main reason for encouraging women to cycle<br>n = 4.1% | Proportion that reported as main reason for encouraging women to cycle<br>n = 3.6% |

| | | | |
|---|---|---|---|
| Transportøkonomisk institutt, 1997 | Mostly middle-aged people with a high level of education and high household income | Proportion that reported as most important improvements to start using a bike | Proportion that reported as most important improvements to start using a bike<br>n = 13% |
| de Cunha, 2018 | Staff and PhD students at the Faculty of Engineering | Proportion that mention circumstances under which they would be more willing to cycle to university | PC=8%<br>C = 41%<br>P=31%<br>A=17% |

*Table 11. Quantitative data for personal enjoyment and improvement-related enablers to cycling for transport*

| **First author, year** | **Population** | **Measure used** | **Improves health and fitness** | **Stress relief** | **Fun and enjoyable activity** | **Improves personal image** |
|---|---|---|---|---|---|---|
| de Geus, 2007 | Working Flemish adults who lived within 10km of their workplace | 5 Point Likert Scale<br>*1 (strongly disagree) - 5 (strongly agree)* | 5 Point Likert Scale<br>*1 (strongly disagree) - 5 (strongly agree)*<br>Mean = 3.96 | | | 5 Point Likert Scale<br>*1 (strongly disagree) - 5 (strongly agree)*<br>Mean = 2.88 |
| Fernández-Heredia, 2014 | 76% students, rest staff of a university in Madrid | 6 Point Likert Scale<br>*1 (not important) to 6 (fundamental)* | 6 Point Likert Scale<br>*1 (not important) to 6 (fundamental)*<br>Mean = 4.89 | | 6 Point Likert Scale<br>*1 (not important) to 6 (fundamental)*<br>Mean = 4.13 | |
| Crawford, 2001 | Two workplaces within Glasgow - one university and one hospital | 5 Point Likert Scale<br>*1 (not important) to 5 (very important)* | | | 5 Point Likert Scale<br>*1 (not important) to 5 (very important)*<br>Mean = 3.6 | |
| Heinen, 2011 | Employees from several large companies in cities in the Netherlands | 5 Point Likert Scale<br>1 (not at all important) to 5 (very important) | 5 Point Likert Scale<br>*1 (not at all important) to 5 (very important)*<br>Mean = 3.38 | 5 Point Likert Scale<br>*1 (not at all important) to 5 (very important)*<br>Mean = 3.82 | | 5 Point Likert Scale<br>*1 (not at all important) to 5 (very important)*<br>Mean = 1.76 |
| Munoz, 2013 | People walking in the centre of Madrid and not tourists. | To what extent to you agree with the following factors in relation to cycling | 10 Point Likert Scale<br>*1 (strongly disagree) to 10 (strongly agree)*<br>Mean = 9.21 | 10 Point Likert Scale<br>*1 (strongly disagree) to 10 (strongly agree)*<br>Mean = 7.04 | 10 Point Likert Scale<br>*1 (strongly disagree) to 10 (strongly agree)*<br>Mean = 8 | 10 Point Likert Scale<br>*1 (strongly disagree) to 10 (strongly agree)*<br>Mean = 7.56 |
| Rérat, 2019 | People who participated in the Bike to Work campaign in Switzerland | Proportion that reported "strongly agree" or "slightly agree" | Proportion that reported "strongly agree" or | Proportion that reported "strongly agree" or | Proportion that reported "strongly agree" or | Proportion that reported "strongly agree" or |

| | | | | | | |
|---|---|---|---|---|---|---|
| | | | "slightly agree"<br>n = 98% | "slightly agree"<br>n = 80% | "slightly agree"<br>n = 88% | "slightly agree"<br>n = 42% |
| Sahlqvist, 2012 | Adult members of Bicycle Queensland | Proportion that reported "strongly agree" or "slightly agree" | Proportion that reported "strongly agree" or "slightly agree"<br>n = 85.8% | | | |
| Swiers, 2017 | Students at an urban UK university | Proportion that reported "strongly agree" or "slightly agree" | Proportion that reported "strongly agree" or "slightly agree"<br>n = 50% | | | |
| Ton, 2019 | Census data from the Netherlands Mobility Panel. Mostly representative of the Dutch population except for teenagers and low-income individuals | 5 Point Likert Scale<br>-2 *(strongly disagree) to 2 (strongly agree)* | | 5 Point Likert Scale<br>-2 *(strongly disagree) to 2 (strongly agree)*<br>Mean = 1.09 | 5 Point Likert Scale<br>-2 *(strongly disagree) to 2 (strongly agree)*<br>Mean = 1.09 | |
| Dill, 2016 | People living in the USA who had ridden a bike in the last 30 days (n = 553) | Proportion that reported as main or somewhat or a reason why they bike | Proportion that reported as:<br>Main reason = 75%<br>Somewhat of a reason = 17% | | Proportion that reported as:<br>Main reason = 65%<br>Somewhat of a reason = 25% | |

*Table 12. Quantitative data for motivation enablers to cycling for transport*

| **First author, year** | **Population** | **Measure used** | **Environmental benefits** |
|---|---|---|---|
| de Geus, 2007 | Working Flemish adults who lived within 10km of their workplace | 5 Point Likert Scale<br>*1 (strongly disagree) - 5 (strongly agree)* | 5 Point Likert Scale<br>*1 (strongly disagree) - 5 (strongly agree)*<br>Mean = 4.21 |
| Fernández-Heredia, 2014 | 76% students, rest staff of a university in Madrid | 6 Point Likert Scale<br>*1 (not important) to 6 (fundamental)* | 6 Point Likert Scale<br>*1 (not important) to 6 (fundamental)*<br>Mean = 5.15 |
| Crawford, 2001 | Two workplaces within Glasgow - one university and one hospital | 5 Point Likert Scale<br>*1 (not important) to 5 (very important)* | 5 Point Likert Scale<br>*1 (not important) to 5 (very important)*<br>Mean = 3.8 |
| Heinen, 2011 | Employees from several large companies in cities in the Netherlands | 5 Point Likert Scale<br>1 (not at all important) to 5 (very important) | 5 Point Likert Scale<br>*1 (not at all important) to 5 (very important)*<br>Mean = 3.62 |
| Munoz, 2013 | People walking in the centre of Madrid and not tourists. | To what extent to you agree with the following factors in relation to cycling | 10 Point Likert Scale<br>*1 (strongly disagree) to 10 (strongly agree)*<br>Mean = 9.75 |
| Rérat, 2019 | People who participated in the Bike to Work campaign in Switzerland | Proportion that reported "strongly agree" or "slightly agree" | Proportion that reported "strongly agree" or "slightly agree"<br>n = 88% |
| Sahlqvist, 2012 | Adult members of Bicycle Queensland | Proportion that reported "strongly agree" or "slightly agree" | Proportion that reported "strongly agree" or "slightly agree"<br>n = 58.3% |
| Swiers, 2017 | Students at an urban UK university | Proportion that reported "strongly agree" or "slightly agree" | Proportion that reported "strongly agree" or "slightly agree"<br>n = 12% |
| Dill, 2016 | People living in the USA who had ridden a bike in the last 30 days (n = 553) | Proportion that reported as main or somewhat or a reason why they bike | Proportion that reported as:<br>Main reason = 18%<br>Somewhat of a reason = 31% |

*Table 13. Quantitative data for practicality-related enablers to cycling for transport*

| **First author, year** | **Population** | **Measure used** | **Efficient** | **Economical transport** | **Flexible** |
|---|---|---|---|---|---|
| Fernández-Heredia, 2014 | 76% students, rest staff of a university in Madrid | 6 Point Likert Scale<br>*1 (not important) to 6 (fundamental)* | 6 Point Likert Scale<br>*1 (not important) to 6 (fundamental)*<br>Mean = 5.08 | 6 Point Likert Scale<br>*1 (not important) to 6 (fundamental)*<br>Mean = 4.77 | 6 Point Likert Scale<br>*1 (not important) to 6 (fundamental)*<br>Mean = 4.87 |
| Crawford, 2001 | Two workplaces within Glasgow - one university and one hospital | 5 Point Likert Scale<br>*1 (not important) to 5 (very important)* | 5 Point Likert Scale<br>*1 (not important) to 5 (very important)*<br>Mean = 3.5 | 5 Point Likert Scale<br>*1 (not important) to 5 (very important)*<br>Mean = 3.5 | |
| Cred Consulting, 2020 | Women living in Sydney, Australia who ride a bike for most of their commute | Proportion that reported as a factor that influenced the route they choose to take | | | Proportion that reported as a factor that influenced the route they choose to take<br>n = 37% |
| Heinen, 2011 | Employees from several large companies in cities in the Netherlands | 5 Point Likert Scale<br>1 (not at all important) to 5 (very important) | 5 Point Likert Scale<br>*1 (not at all important) to 5 (very important)*<br>Mean = 4.16 | 5 Point Likert Scale<br>*1 (not at all important) to 5 (very important)*<br>Mean = 3.74 | 5 Point Likert Scale<br>*1 (not at all important) to 5 (very important)*<br>Mean = 4.2 |
| Munoz, 2013 | People walking in the centre of Madrid and not tourists. | To what extent to you agree with the following factors in relation to cycling | 10 Point Likert Scale<br>*1 (strongly disagree) to 10 (strongly agree)*<br>Mean = 7.67 | 10 Point Likert Scale<br>*1 (strongly disagree) to 10 (strongly agree)*<br>Mean = 9.16 | 10 Point Likert Scale<br>*1 (strongly disagree) to 10 (strongly agree)*<br>Mean = 8.41 |
| Rérat, 2019 | People who participated in the Bike to Work campaign in Switzerland | Proportion that reported "strongly agree" or "slightly agree" | Proportion that reported "strongly agree" or "slightly agree"<br>n = 60% | Proportion that reported "strongly agree" or "slightly agree"<br>n = 53% | Proportion that reported "strongly agree" or "slightly agree"<br>n = 90% |
| Sahlqvist, 2012 | Adult members of Bicycle Queensland | Proportion that reported "strongly agree" or "slightly agree" | Proportion that reported "strongly agree" or "slightly agree"<br>n = 64.6% | | |
| Swiers, 2017 | Students at an urban UK university | Proportion that reported "strongly agree" or "slightly agree" | Proportion that reported "strongly agree" or "slightly agree"<br>n = 46% | | |
| Winters, 2011 | Current and potential cyclists in Vancouver from a random sample of home addresses | 3 Point Likert Scale<br>*-1 (much less likely to cycle) to 1 (much more likely to cycle)* | 3 Point Likert Scale<br>*-1 (much less likely to cycle) to 1 (much more likely to cycle)*<br>Mean - 0.53 | | |
| Ton, 2019 | Census data from the Netherlands Mobility Panel. Mostly representative of the | 5 Point Likert Scale<br>-2 *(strongly disagree) to 2 (strongly agree)* | 5 Point Likert Scale<br>-2 *(strongly disagree) to 2* | | 5 Point Likert Scale<br>-2 *(strongly disagree) to 2* |

| | | | | | |
|---|---|---|---|---|---|
| | Dutch population except for teenagers and low-income individuals | | *(strongly agree)*<br>Mean = 0.6 | | *(strongly agree)*<br>Mean = 1.29 |
| Dill, 2016 | People living in the USA who had ridden a bike in the last 30 days (n = 553) | Proportion that reported as main or somewhat or a reason why they bike | Proportion that reported as:<br>Main reason = 8%<br>Somewhat of a reason = 18% | Proportion that reported as:<br>Main reason = 17%<br>Somewhat of a reason = 27% | Proportion that reported as:<br>Main reason = 11%<br>Somewhat of a reason = 21% |

#### Supplementary Material D - Search Strategy

The research question and search terms were guided by the PICO framework (Population, Intervention, Comparison and Outcome). As no comparison or population was specified for the search, these sections of the PICO framework were excluded. Other search terms using the ‘adj’ function to identify topical research has been indicated as ‘other search terms’. Each section of the framework was combined with the ‘AND’ function. A summary of the terms used across databases is shown below.

*Table 27. Summary of search terms*

| | Terms |
|---|---|
| Intervention | Bicycl* OR cycl* OR biking* OR bike* OR commut* OR active commute OR active transport* OR active travel* OR |
| Outcome | Barrier* OR prevent* OR attitud* OR knowledge OR behavi?r* OR enable* OR motivat* OR facilitat* OR |
| Other search terms | ((bicycl* or cycl* or bike* or biking*) adj4 (attitude* or enable* or barrier* or motivat* or facilitat* or prevent*)) |
